# Gut microbiota link dietary fiber intake and short-chain fatty acid metabolism with eating behaviour

**DOI:** 10.1101/2021.02.18.21251818

**Authors:** Evelyn Medawar, Sven-Bastiaan Haange, Ulrike Rolle-Kampczyk, Beatrice Engelmann, Arne Dietrich, Ronja Thieleking, Charlotte Wiegank, Charlotte Fries, Annette Horstmann, Arno Villringer, Martin von Bergen, Wiebke Fenske, A. Veronica Witte

## Abstract

**Background:** The gut microbiome modulates human brain function and eating behavior through multiple factors, including short-chain fatty acid (SCFA) signaling. We aimed to explore which bacterial genera relate to eating behavior, diet and SCFA metabolites in overweight adults. In addition, we tested whether eating-related microbiota relate to treatment success in patients after Roux-en-Y gastric bypass (RYGB).

**Methods:** Anthropometrics, dietary fibre intake, eating behaviour, 16S-rRNA-derived microbiota and SCFA measured in feces and blood were correlated in young overweight adults (n=27 (9F), 21-36 years, BMI 25-31 kg/m2). Correlated genera were compared in RYGB (n=23 (16F), 41-70 years, BMI 25-62 kg/m2) and in control patients (n=17 (11F), 26-69 years, BMI 25-48 kg/m2).

**Results:** In young adults, 7 bacteria genera, i.e. Alistipes, Blautia, Clostridiales cluster XVIII, Gemmiger, Roseburia, Ruminococcus and Streptococcus, correlated with healthier eating behavior, while 5 genera, i.e. Clostridiales cluster IV and XIVb, Collinsella, Fusicatenibacter and Parabacteroides, correlated with unhealthier eating (all |r| > 0.4, FDR-corrected p < 0.05). Some of these genera including Parabacteroides related to fiber intake and SCFA metabolites, and to weight status and treatment response in overweight/obese patients.

**Conclusions:** Specific bacterial genera, particularly Parabacteroides, were associated with weight status and eating behavior in two independent well-characterized cross-sectional samples. These exploratory findings indicate two groups of presumably beneficial and unfavourable genera that relate to eating behaviour and weight status, and indicate that dietary fiber and SCFA metabolism may modify these relationships.

## Background

Gut microbes modulate brain function and behavior via immune, endocrine, neural and humoral routes [1]. This could play a key role in neuronal feeding circuits and overeating, as dysbiosis of the microbiota composition has been documented in psychiatric eating disorders [2] and obesity [3].

However, nutrition- or body weight-related microbial changes and their functional relevance are still relatively unclear. In mice, gastric bypass-related differences in the microbiota profile, such as a higher abundance of the genera *Escherichia (*phylum *Proteobacteria)* and *Akkermansia (*phylum *Verrucomicrobia)*, induced weight loss when transferred to germ-free animals [4]. In humans, bariatric surgery similarly led to higher overall microbiota diversity and to higher abundance of the species *Escherichia coli* and in some studies to further abundance changes within the phylum *Bacteroidetes*, such as a higher post-surgery ratio of the genera *Bacteroides to Prevotella* [5] and less *Firmicutes* (phylum level) or to more *Gammaproteobacteria* (class level) [6]. The ratio of *Bacteroides to Prevotella* at baseline predicted dietary weight loss success after 24 weeks in an intervention study in 80 overweight individuals [7]. Further, a one-week dietary intervention trial in 20 individuals found that microbial composition predicted glycemic response [8].

Human-to-mouse fecal transplant experiments further underline the causal role of specific microbiota to facilitate weight loss [9], and human-to-human microbiota transplantation experiments increased insulin sensitivity according to [10]. In a recent human study, accompanied by mouse model data, an individual’s microbiota profile, extracted from fecal samples during periods of dietary weight loss, prevented weight regain when transferred back to the individuum orally [11].

Mechanistic insights into how specific gut bacteria modulate human eating behaviour and weight status are still limited. The gut microbiota is supposed to affect the host’s metabolism by altering energy extraction from food, and by modulating dietary or host-derived compounds that modify the metabolic pathways of the host [12]. For example, short-chain fatty acids (SCFA) are excreted by certain gut bacteria as a result of carbohydrate fermentation, and SCFA stimulate the secretion of anorexigenic hormones, such as peptide YY (peptide tyrosine tyrosine or PYY) and glucagon-like-peptide-1 (GLP-1) in the colon, which further signal to hypothalamic nuclei as one mechanism of homeostatic regulation [13]. SCFA can also cross the blood-brain-barrier and act as signaling molecules in the brain to directly modulate appetite and food-decision making [1]. First interventional studies showed that intake of butyrate (one type of SCFA) or the butyrate-producing bacteria *Akkermansia spp*. exert beneficial effects on body weight depending on treatment intention in humans [14] and on brain functions in mice [15], including reduced food intake [16]. Notably, specific pre-biotic nutrients, such as dietary fibers, are known to nourish SCFA-producing bacteria in the gut, rendering diet a potent modifier of gut-brain signalling [17].

In sum, the gut microbiome may influence feeding behaviour, e.g. by modulating reward and homeostatic signaling [18, 19] and by stimulating the vagal nerve [20], in particular in dysregulated biological systems, such as in food addiction [21] or eating disorders [2]. Yet, direct knowledge if specific genera are linked to eating behavior via dietary intake and SCFA in humans is lacking. Here, we asked whether gut microbial diversity and genera abundance relate to eating behaviour, and to SCFA metabolites in the colon (feces) and in the periphery (blood) in a homogenous sample of young overweight adults. In addition, we tested whether the abundance of microbiota that related to eating behavior in that overweight sample correlate with weight status, eating behaviour and treatment success (i.e. achieved weight loss) in another sample, i.e. patients at two years after bariatric surgery and control overweight/obese patients.

## Methods

### Samples characteristics and data collection

We included all participants with available microbiota datasets measured at a cross-sectional timepoint from two studies. Sample 1 comprised of 27 healthy young overweight adults (9F, 21-36 years, BMI 25-31 kg/m^2^) drawn from a randomized clinical trial (Clinical Trials registration NCT03829189), where baseline data was available from ongoing data collection until January 2021. Sample 2 comprised of 23 patients two years after Roux-en-Y gastric bypass (RYGB) surgery (see below; “good responders”: n = 11 (7F), 41-70 years, BMI 25-29 kg/m^2^; “bad responders”: n = 12 (9F), 31-67 years, BMI 41-62 kg/m^2^), as well as age-, gender- and BMI-matched controls (overweight: n = 8 (5F), 41-58 years, BMI 25-29 kg/m^2^; obese: n = 9 (6F), 26-70 years, BMI 41-48 kg/m^2^), drawn from an observational study where data collection was completed (ethics proposal 027/17-ek).

All participants donated feces (see **SI**) for microbiota analysis (Shannon effective [22] and relative abundance of microbiota genera), underwent anthropometric measurements and filled in questionnaires to quantify eating behaviour traits. Also, data on dietary fibre intake, hunger ratings after a standardized meal and SCFA in blood and feces were available in sample 1 (see below).

### Microbiota assessment

To assess microbiota community structure we used 16S rRNA gene profiling of the fecal samples. Therefore, DNA was extracted and V3-V4 variable regions of the 16S rRNA genes were amplified by PCR and a library was constructed, followed by paired-end 2×250bp Illumina sequencing. These analyses were done by GENEWIZ Germany GmbH, Leipzig. Next, the inhouse Galaxy server using a pipeline implemented with the DADA2 R package processed raw data in fastq format. For each sample, paired-end reads were joined, low-quality reads were removed, reads were corrected, chimeras removed and Amplicon Sequence Variants (ASVs) were obtained. Taxonomy was annotated to the ASVs using the RDP database [23]. The read counts per ASV with taxonomic annotation were normalized and relative abundances of each ASV and taxa were calculated using the R scripts Rhea. Visualization of all library-indexed genera was done as in [24] by inhouse written R-tools using ggplot2.

### Eating behaviour

To characterise eating behaviour traits, questionnaires based on self-report were used: the Three-Factor Eating Questionnaire (TFE-Q) (German version, [25]) and the Eating Disorder Examination Questionnaire (EDE-Q) (German version, [26]) as available for sample 1 and sample 2, respectively. The TFE-Q assesses three domains of eating behaviour (cognitive restraint, disinhibition, hunger), and the EDE-Q covers the subscales dietary restraint, eating concern, weight concern, and shape concern. Scoring was performed according to the respective manuals.

#### Additional analyses in sample 1

From all measurements available in sample 1 in the context of the RCT (see above), we additionally considered all available hunger ratings after a standardized meal (3 out of 4 measures, 1 with missing data) and all available dietary fiber intake data (from a quantitative food frequency questionnaire, fiber in g/day and fiber per 1000 kcal). We further considered anthropometric assessments to be of interest in this study and limited those to two major health indicators, i.e. systolic blood pressure (mean of three consecutive measurements) and relative body fat (%) obtained from bioelectrical impedance analysis **(see SI)**. Blood was obtained in fasting state (12 ± 3h fasted) and samples were centrifuged at 3500 revolutions per minute at 7°C for 6 minutes. Serum was aliquoted within one hour of obtainment. Processed aliquots were stored at −80°C until data analysis. For SCFA in blood and stool, analysed according to [27], we focused on three major SCFAs out of eight measured, i.e. acetate, butyrate, and propionate (see SI). All other measures were not considered of interest to the current analyses.

#### Obesity surgery in sample 2

For sample 2, RYGB (**see SI**) patients were selected for microbiota analysis based on their response to the surgical treatment. Specifically, RYGB patients were identified from the database of the University of Leipzig if their surgery dated back at least 2 years and all those were further divided in percentiles according to pre-defined relative excessive weight loss (EWL) thresholds, resulting in 23 RYGB patients good responders: sustained EWL > 70 %, mean 93% ± 4 SD, range 86-98%, n = 12; bad responders: sustained EWL < 40 %, mean 20% ± 13 SD, range 3-35%, n = 11). Next, obese and overweight control patients were selected from the database based on age, sex and BMI to match those two groups of RYGB patients. Afterwards, RYGB patients only filled in a series of questionnaires, performed cognitive tests and donated blood for another study purpose; and fecal samples of all patients were analysed. From this dataset, we considered of interest to the current analysis the following variables: weight loss after surgery (in kg and in BMI), all available eating questionnaire data (3 EDEQ scales, see above) and those microbiota genera abundance based on 16S rRNA sequencing.

### Statistical analysis

#### Correlational analysis

Relative taxa abundance (%) on the genera level was used as primary variables of interest. Non-normally distributed variables were log- or Tukey-transformed, so that skewness of < |1| was reached (**for details Suppl. Fig. 2**). No observations were eliminated, instead all microbiota-cases were complete and included. For the main analysis, 20 out of 121 genera were included as they appeared in at least 80% of individuals [30] and fed into a correlation matrix with all variables of interest in sample 1 (37 variables in total, see above), i.e. Shannon index, 3 TFEQ traits, 3 hunger ratings, body fat, systolic blood pressure, dietary fiber intake (g/day and g/1000kcal) and 3 SCFA each in feces and blood, respectively. All values were FDR-corrected and statistical significance was set to p < 0.05. Those genera that were significantly associated with eating behavior (TFEQ traits and/or hunger ratings, p-FDR < 0.05) were then correlated with weight status and RYGB treatment success in sample 2. Group differences across overweight, obese, good and bad RYGB responders were tested with non-parametric Kruskal-Wallis tests. Further correlations were tested with Pearson’s correlation coefficient r for normally distributed variables or with Spearman’s rho for non-normally distributed variables. Explorative analysis considerations were addressed according to [28] (**see SI**).

To further investigate, if the interplay of correlated genera - rather as a holobiont than individually - is determinative of the observed relations, the relation between correlated to non-correlated genera was computed by three composite scores (1)-(3).

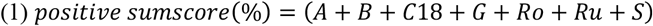

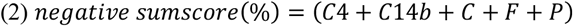

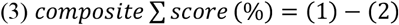

#### Mediation Analysis

Using simple mediation analysis using medmod (https://cran.r-project.org/web/packages/medmod/index.html) in RStudio version 3.6.1, we checked for statistical mediation in sample 1 for variables showing bivariate correlations in the following paths:

i. fiber —> correlated genera or sumscores —> eating behaviour (TFEQ, hunger ratings)
ii. eating behaviour (TFEQ, hunger ratings) —> fiber —> correlated genera or sumscores
iii. correlated genera or sumscores —> SCFA —> eating behaviour (TFEQ, hunger ratings)

Significance was set to p < 0.05, and the main analysis for sample 1 was corrected for multiple testing using the false-detection rate (FDR)-correction. All analyses were performed in RStudio version 3.6.1.

## Results

Characteristics of sample 1 and 2 are listed below (see **Table 1-2**). Eating behaviour traits varied across both samples, and in sample 2, restrained eating and shape/weight concerns differed between those that achieved long-term excessive weight loss after bariatric surgery compared to those that did not (good vs. bad responders, all W> 58.5, p < 0.001, **Tab. 2, Suppl. Figures 1**).

**Table 1:** Descriptives for sample 1.

|  | *Sample 1* |  |  |  |
| --- | --- | --- | --- | --- |
| *n, sex (F/M)* | *27 (9F/18 M)* |  |  |  |
|  | *mean* | *SD* | *minimum* | *maximum* |
| *age (years)* | 28.4 | 4.5 | 21 | 36 |
| *education (SES index)*<br>*(score from 3 to 21)*<br>*(four NAs)* | 15.0 | 2.8 | 8.2 | 19.2 |
| *BMI (kg/m<sup>2</sup>)* | 27.7 | 1.7 | 25.0 | 31.2 |
| *TFEQ cognitive restraint (sum score)* | 5.6 | 4.1 | 0 | 13 |
| *TFEQ disinhibition (sum score)* | 6.0 | 2.1 | 2 | 11 |
| *TFEQ hunger (sum score)* | 5.0 | 3.4 | 0 | 12 |
| *time fasted (h)* | 12.5 | 2.7 | 6 | 18 |
| *hunger 15min postprandial (1-8 scale)* | 4.2 | 1.7 | 1 | 7 |
| *hunger 40min postprandial (1-8 scale)* | 5.3 | 1.3 | 2 | 7 |
| *hunger 65min postprandial (1-8 scale)* | 5.3 | 1.4 | 2 | 8 |
| *mean systolic blood pressure (mmHg)* | 128.0 | 10.9 | 107.0 | 152.7 |
| *% fat mass (female, male)* | 34.7 (F)<br>22.8 (M) | 4.2 (F)<br>5.2 (M) | 27.3 (F)<br>7.6 (M) | 39.8 (F)<br>30.7 (M) |
| *habitual fiber intake / 1000 kcal / d (g)* | 10.4 | 3.6 | 4.4 | 20.0 |

**Table 2:**
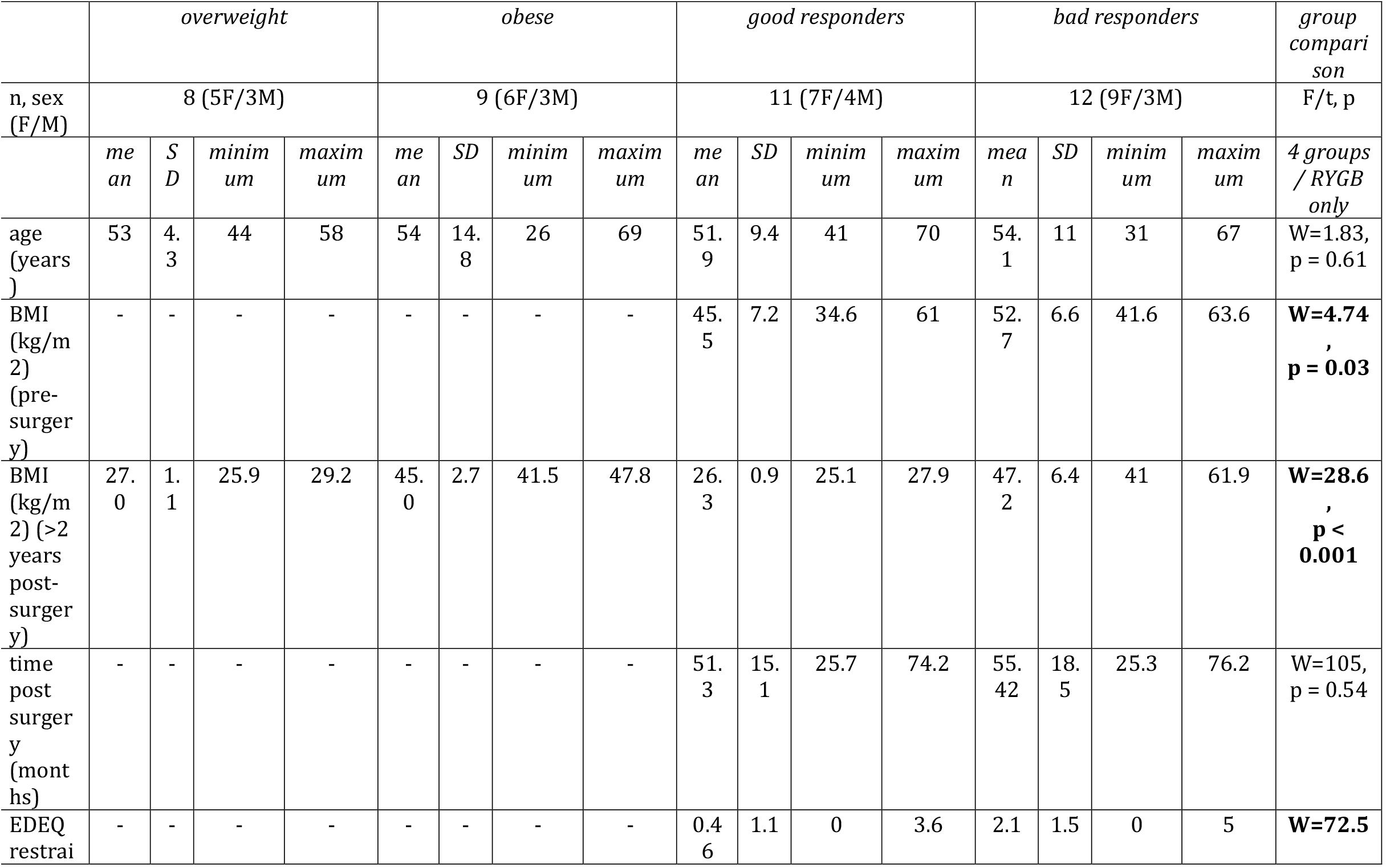
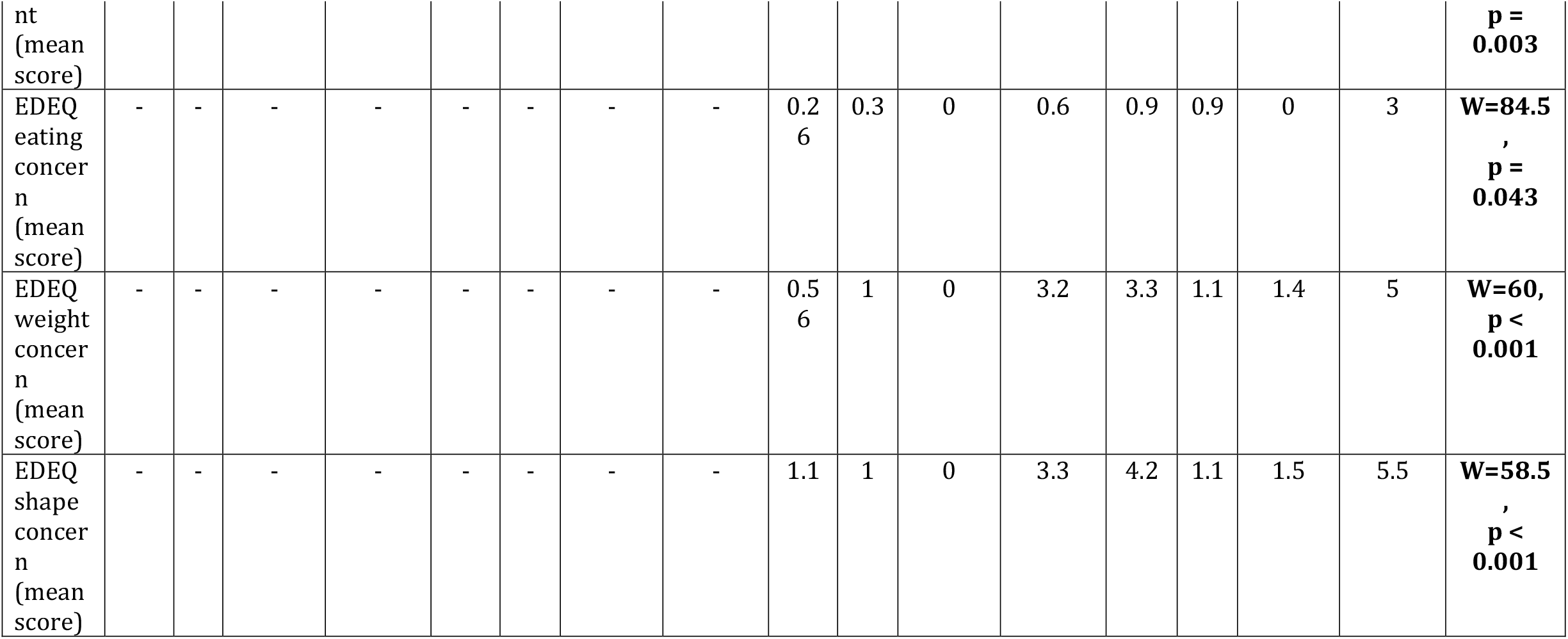
Descriptives for sample 2.

Overall microbiota diversity at the phylum level was relatively comparable across participants of samples 1 and 2 except higher ratio of Firmicutes to Bacteroidetes in sample 1, and Prevotellaceae and Fusobacteriaceae families were more abundant in patients after RYGB surgery (**Figure 1, Suppl Fig. 3-4**, see **SI** for details).

**Figure 1.**
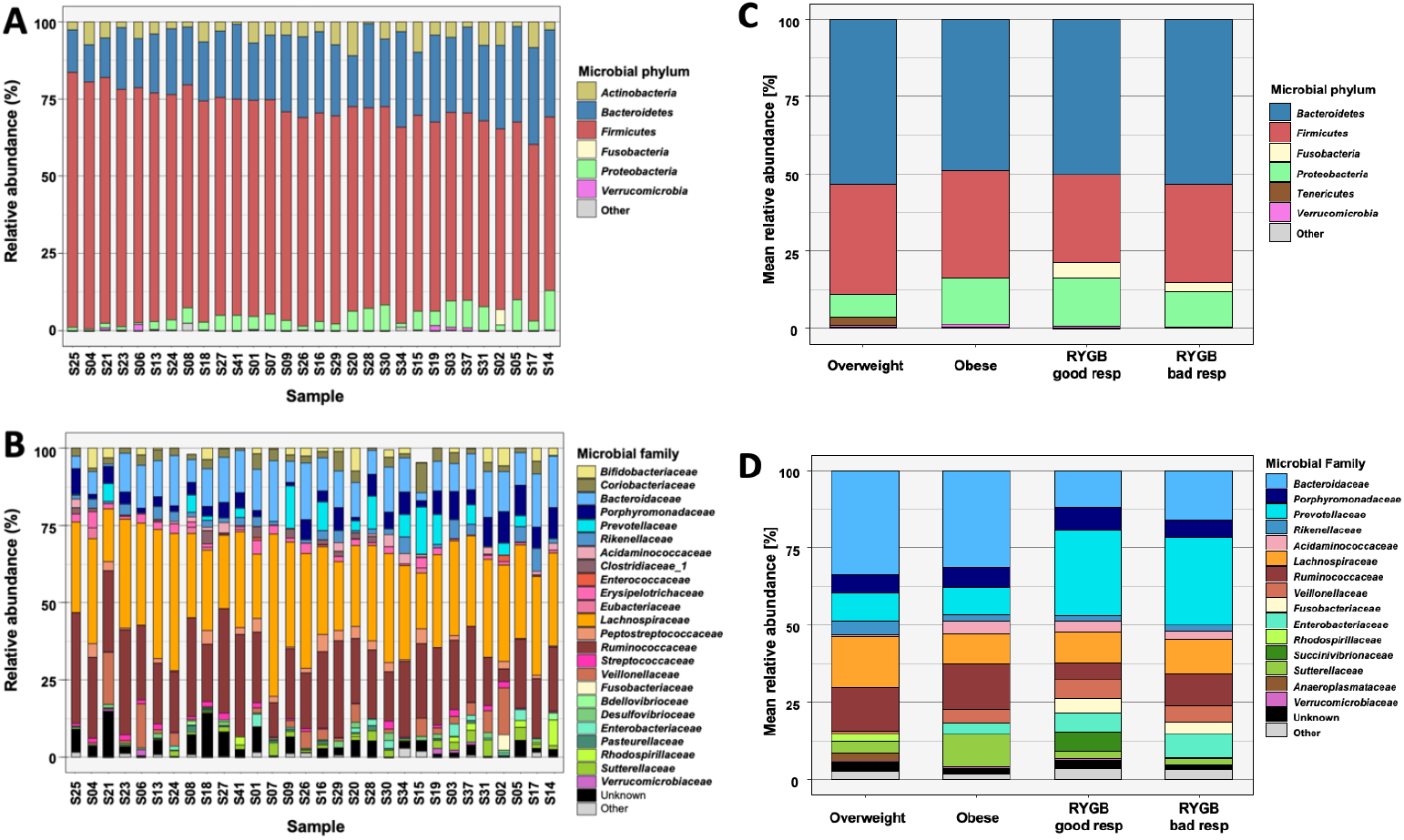
Relative abundance per subject or per group on microbial phylum (A,C) and family level (B,D) (sorted by Firmicutes abundance).

### Microbiota, eating behaviour traits and health indicators in overweight adults

In sample 1, higher relative abundance of *Collinsella* (phylum Actinobacteria), *Clostridium* IV and XIVb, *Fusicatenibacter* (all three phylum Firmicutes) and *Parabacteroides* (phylum Bacteroidetes) were related to less healthy eating behaviour (higher TFEQ scores and/or higher hunger ratings, all 0.61 < |r| > 0.42, p-FDR < 0.05, **Figure 2A**). Contrastingly, higher relative abundance of the microbial genera *Alistipes* (phylum Bacteroidetes), *Blautia, Clostridium* XVIII, *Gemmiger, Roseburia, Ruminococcus* and *Streptococcus* (all phylum Firmicutes) correlated with healthier eating behaviour (all 0.76 < |r| > 0.42, p-FDR < 0.05, **Figure 2B, Suppl. Figure 5**).

**Figure 2.**
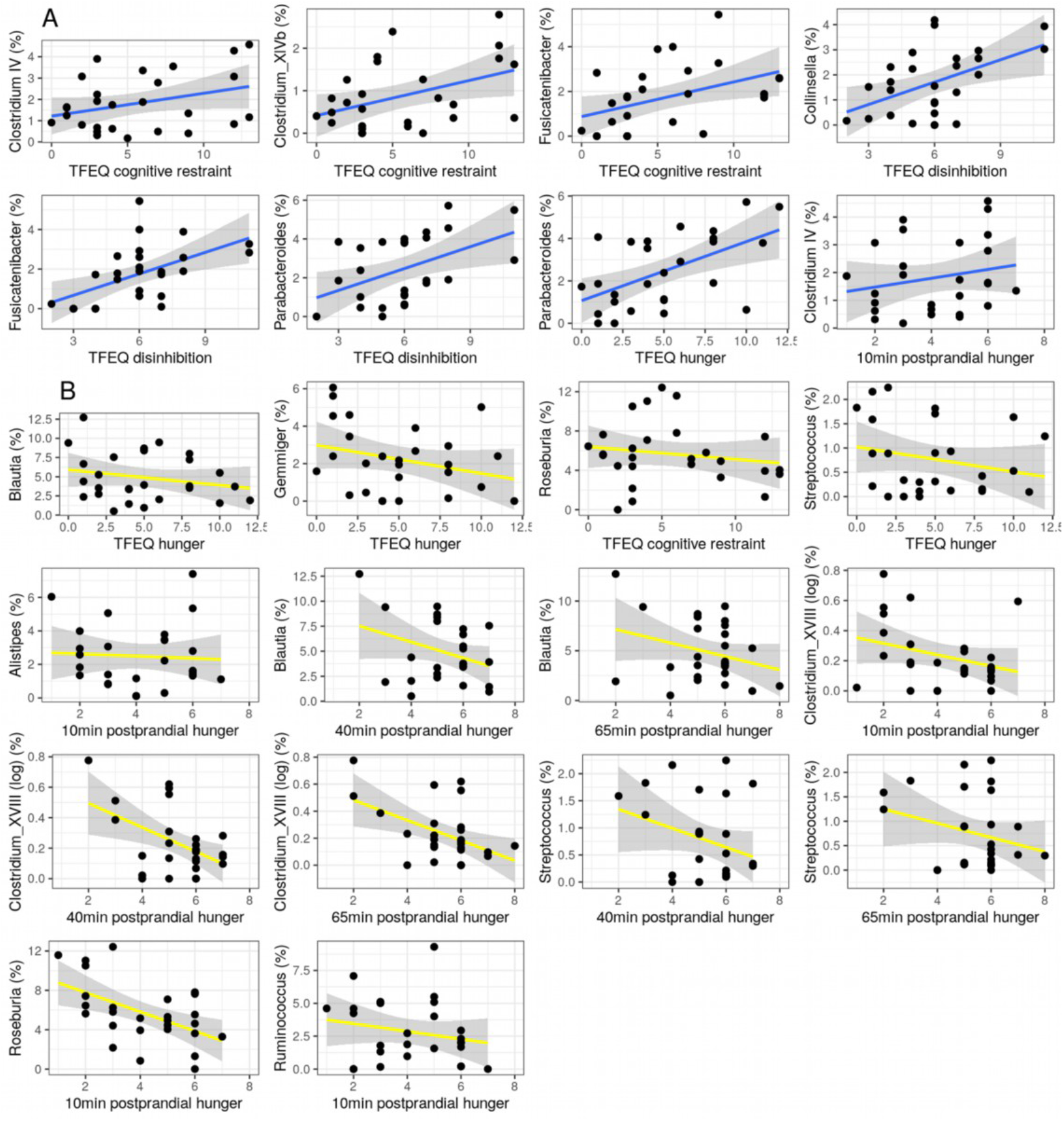
Correlations between eating behaviour traits (TFEQ and hunger ratings) or health indicators and bacterial genera in overweight adults (all |r| > 0.42, all p-FDR < 0.05; sample 1). A: inversely health-related genera; B: health-related genera. C: Collinsella and body fat mass; D: Streptococcus and mean systolic blood pressure.

Further, *Collinsella* abundance significantly correlated with higher body fat mass (sex-standardized, r = 0.61, p < 0.001, **Figure 2C**). *Streptococcus* abundance was significantly correlated with lower mean systolic blood pressure (r = - 0.70, p-FDR < 0.001, **Figure 2D**).

### Relation to dietary fiber intake and SCFA

Out of the 12 genera that were significantly associated with eating behaviour (from now on called “(inversely) health-related” genera), three were associated with lower (*Collinsella* and *Parabacteroides*) or higher (*Clostridium XVIII*) dietary fiber intake (all 0.73 < |r| > 0.49, p-FDR < 0.05, **Figure 3A-C**). Moreover, higher dietary fiber intake per se was significantly associated with lower disinhibited eating (r = - 0.58, p-FDR < 0.01) and lower body fat mass (r = - 0.75, p-FDR < 0.0001, **Figure 3D-E**).

**Figure 3.**
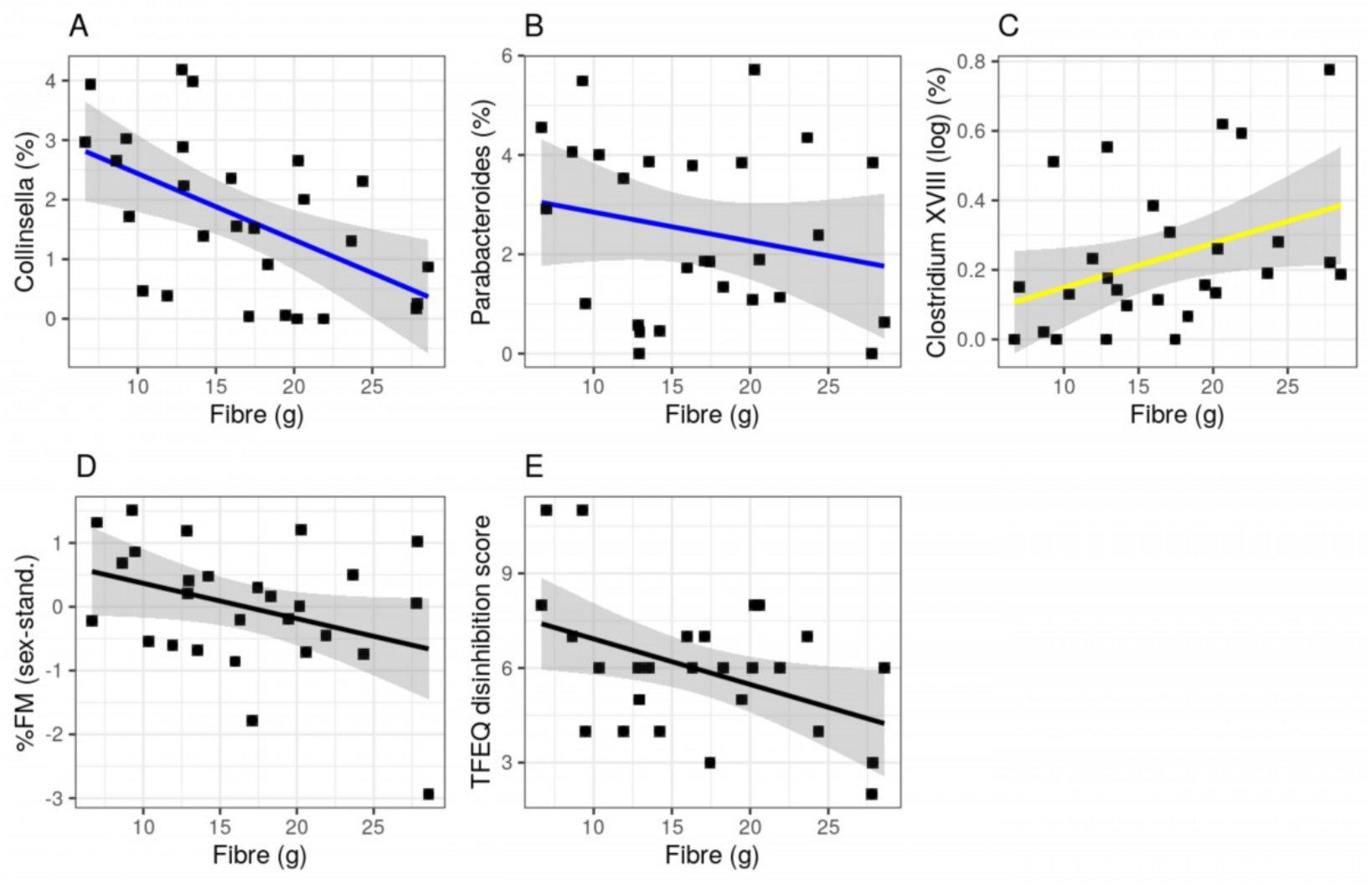
Associations between fiber intake and inversely health-related genera (A-B), health-related genera (C), body fat mass (D) and eating trait disinhibition (E) (Pearson’s correlation all 0.75 < |r| > 0.58, all p-FDR < 0.05; sample 1 n=27).

SCFA concentrations in feces were highly variable and up to ∼1000 times higher compared to serum for all three measured SCFA (all t(24) > 11.6, p < 0.001). Serum acetate was 2.5 times higher compared to butyrate and propionate in serum (**Suppl. Table 1**).

We observed that higher abundance of some of the inversely health-related genera correlated with higher levels of different SCFA in feces and serum (all r > 0.50, p-FDR < 0.01). In addition, most health-related genera correlated with some feces and serum SCFA markers, however revealing both positive and negative associations (only those associated with eating behaviour were considered, all 0.65 < |r| > 0.44, p-FDR < 0.05, **Suppl. Figure 6**). Note, that some genera showed differential correlations within the different SCFA, e.g. higher *Alistipes* correlated with higher acetate in both feces and serum and with fecal butyrate, but with lower fecal propionate. Moreover, considering the inversely correlated genera, *Fusicatenibacter* and *Parabacteroides* correlated significantly with higher fecal concentrations of propionate and acetate, respectively.

Also, higher fecal propionate levels correlated significantly with higher cognitive restraint eating (r = 0.50, p-FDR = 0.014, **Figure 4A**). Higher fecal acetate, butyrate and propionate levels correlated with higher hunger ratings (all r > 0.45, all p-FDR < 0.04), but also serum propionate with hunger (r = 0.45, p-FDR = 0.03). Moreover, serum acetate and butyrate were inversely associated with body fat mass (all r > - 0.43, all p-FDR < 0.04) (**Figure 4B-C**). Notably, serum levels did not correlate with fecal SCFA concentrations (all r < | 0.17 |, all p-uncorr < 0.86, **Suppl. Figure 7**).

**Figure 4.**
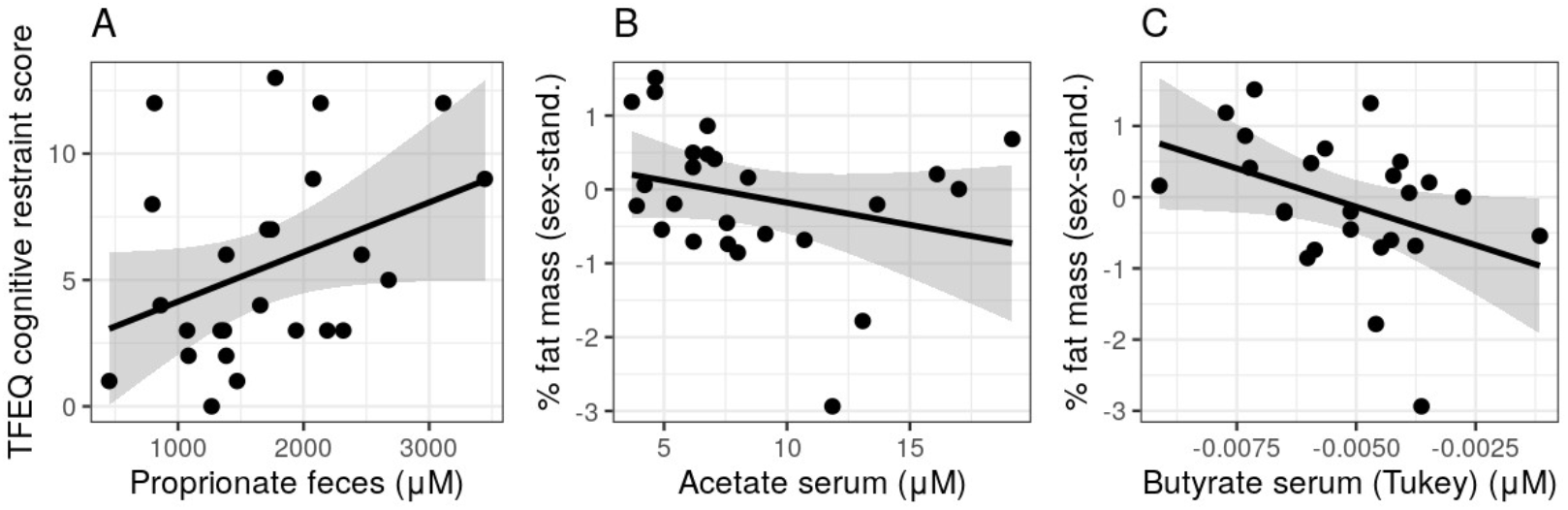
Associations of SCFA levels in feces and eating traits (A) and serum (B-C) and body fat mass.

### Genera sum score and mediation analyses

The negative sumscore of the five inversely health-related genera abundance resulted in significant correlations for two of the eating traits (cognitive restraint r = 0.59, p-uncorr = 0.001; disinhibition r = 0.65, p-uncorr < 0.001, **Figure 5A-B**). The positive sumscore of the seven health-related genera showed no significant associations (all p-uncorr < 0.95, **Suppl. Figure 9**). Neither sumscore correlated with fecal or serum SCFA levels.

**Figure 5.**
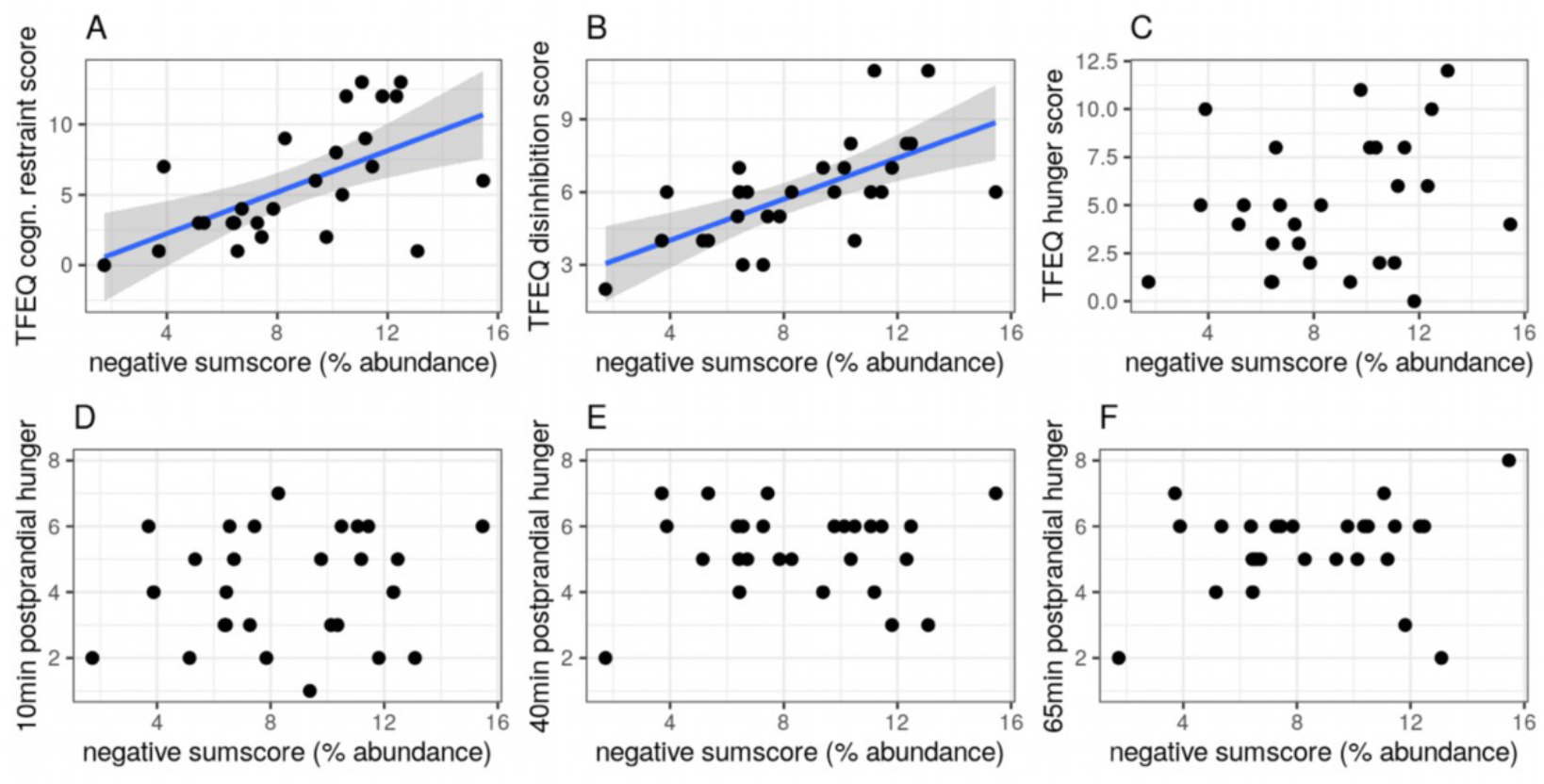
Correlations between microbial genera sums score for inversely health-related genera with respect to eating behaviour outcomes.

Exploratory mediation path analyses of the proposed models did not show statistically significant mediating paths for differences in diet, eating behaviour or hunger ratings through differences in *Parabacteroides* or positive/negative sumscores (**Suppl. Table 2-3**). Considering SCFA, similar results emerged, except for acetate: Here, while the direct effect c’ did not reach significance (ß = −0.3, p = 0.13), higher *Parabacteroides* abundance was linked with higher postprandial hunger ratings through higher fecal acetate levels (indirect effect, a*b, ß = 0.36, 95% CI [0.05 0.66], p = 0.02, **Suppl. Table 4**).

### Mircobiota genera differences between overweight, obese and surgery groups

In sample 2, we aimed to confirm links between the genera of interest from sample 1 and treatment success and eating behaviour. Two of the five inversely health-related genera were significantly different between groups (all H(3) > 9.5, p < 0.023) with lower relative abundance of *Parabacteroides* in good vs. bad responders (H(1) = 4.9, p = 0.027) **Figure 6**). In addition, six of the seven health-related genera were more abundant in the overweight group (all H(3) > 8.3, p < 0.036, **Figure 7**), but did not differ in the good vs. bad RYGB responders.

**Figure 6.**
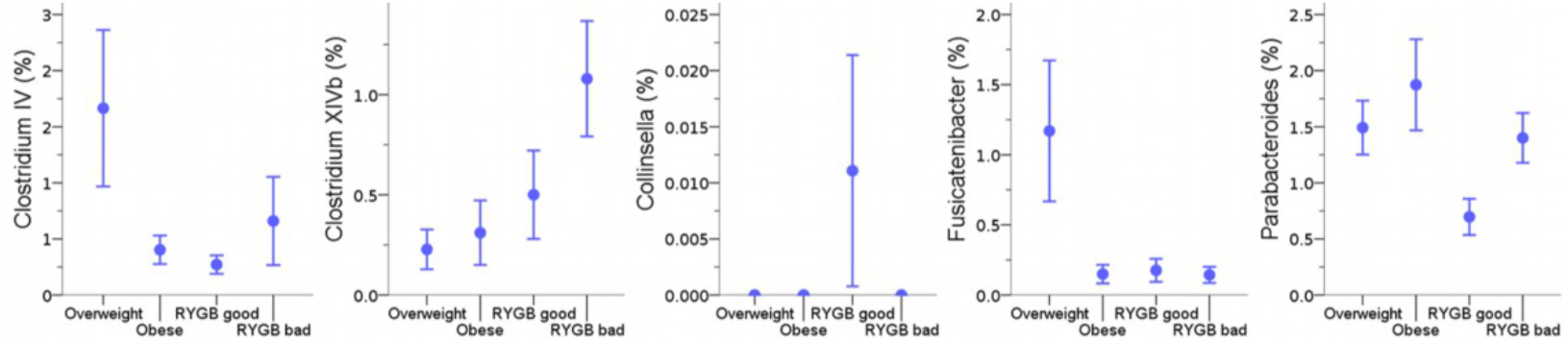
Differences in relative bacterial abundance across groups in sample 2 for inversely health-related genera detected in sample 1.

**Figure 7.**
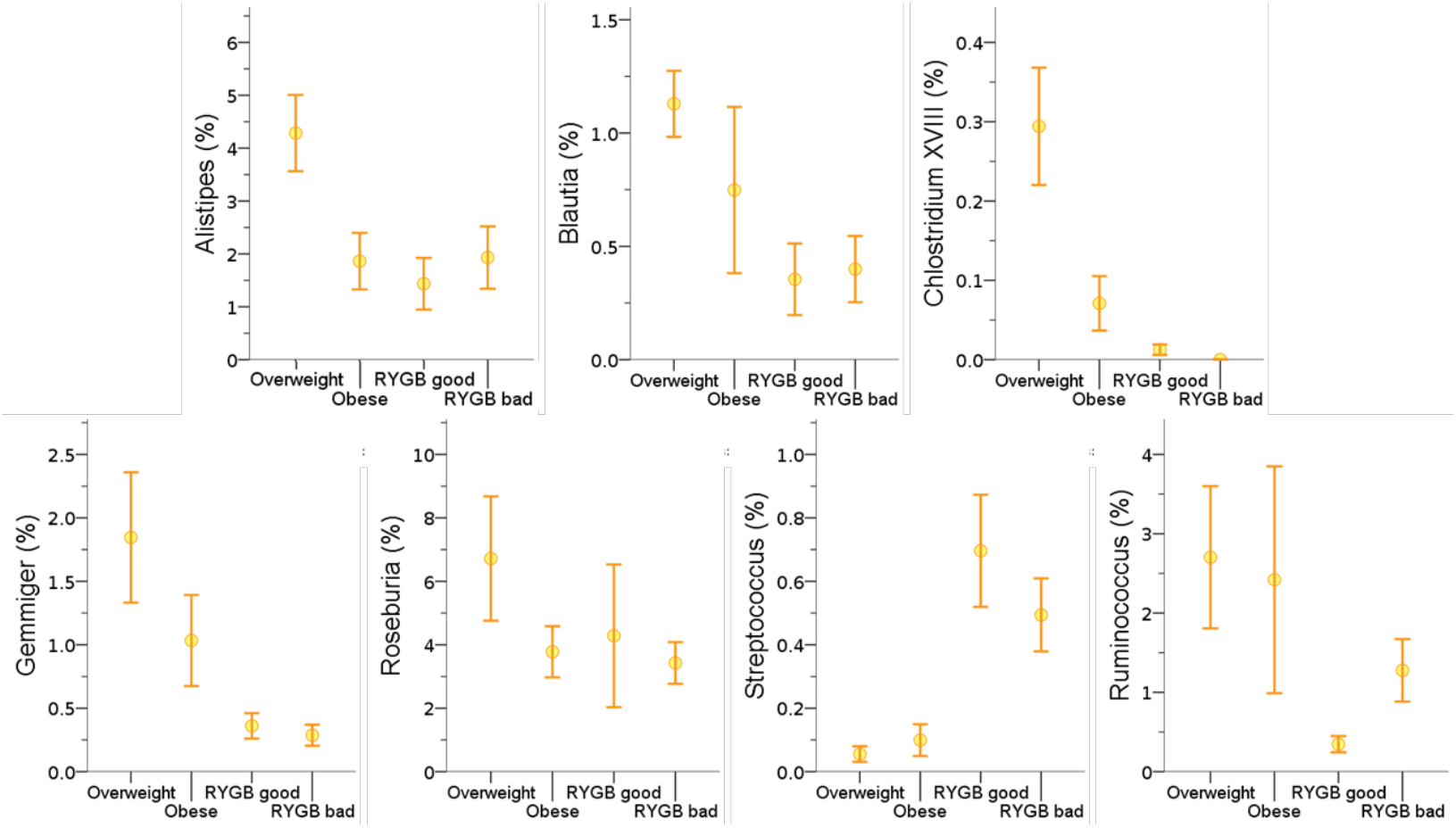
Differences in relative bacterial abundance across groups in sample 2 for positively-related genera detected in sample 1.

Considering the sumscores, we found that both sumscores differed between groups (**Figure 8**, all H(3) > 11.3, p < 0.01) with the negative sumscore showing higher values in the bad vs. good RYGB responders (H(1) = 2.1, p = 0.036). In addition, both the positive (n.s.; H(1) = 1.9, p = 0.05) and the negative sumscore (H(1) = 2.02, p = 0.043) showed higher values in overweight vs. obese participants.

**Figure 8.**
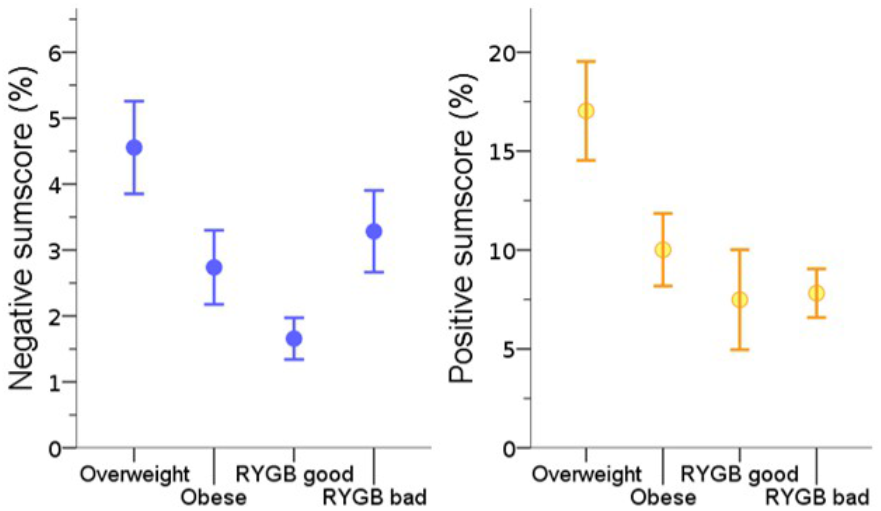
Differences in relative bacterial abundance across groups in sample 2 for positive and negative sumscores of all related genera detected in sample 1.

Bad vs. good RYGB responders showed higher eating restraint scores (H = 5.3, p = 0.022, **Suppl. Figure 1**), and higher scores correlated with higher *Parabacteroides* abundance in these groups (r = 0.44, p = 0.039, **Figure 9A**). Moreover, lower *Parabacteroides* abundance correlated significantly with higher weight loss after surgery (r = 0.49, p = 0.019, n = 20, **Figure 9B**). The negative sumscore correlated with unhealthier eating behaviour (**Figure 9C**, mean of all EDEQ subscales, r = 0.47, p = 0.027; EDEQ restraint, r = 0.49, p = 0.022) and with less weight loss after surgery (**Figure 9D**, weight, r = 0.53, p = 0.011, BMI, r = 0.53, p = 0.011).

**Figure 9.**
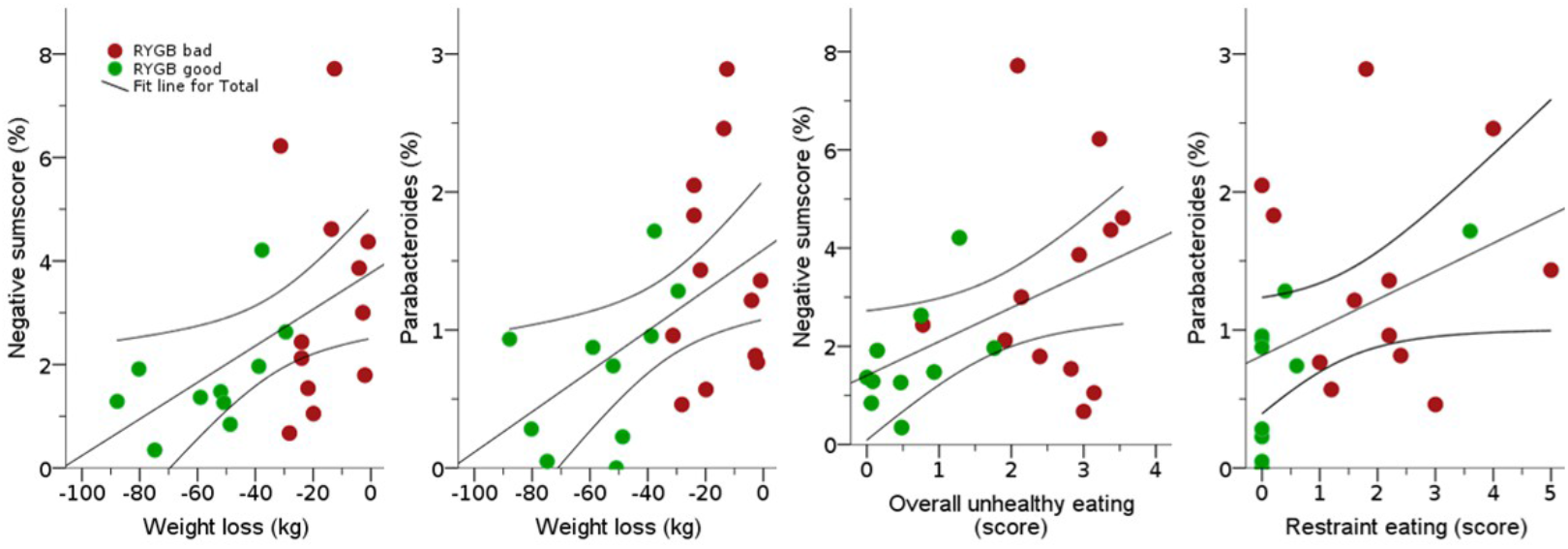
Negative genera sumscore and *Parabacteroides* abundance link to unhealthier eating behavior and less weight loss after RYGB surgery. RYGB, Roux-en-Y gastric bypass.

## Discussion

Combining data from two human cross-sectional datasets, this exploratory analysis finds two groups of microbiota genera that were either positively or inversely associated with both healthier eating behaviour and anthropometrics (1) in a deeply phenotyped sample of young overweight adults and (2) when comparing microbiota observed in (1) in patients showing a good or bad response two years after bariatric surgery with matched controls, respectively. More specifically, in young overweight adults, 7 bacterial genera, i.e. *Alistipes, Blautia, Clostridium XVIII, Gemmiger, Roseburia, Ruminococcus* and *Streptococcus*, correlated with healthier eating behaviour traits and lower subjective hunger ratings, indicating potential benefits for the host metabolism, while 5 bacterial genera, i.e. *Clostridum IV, Clostridium XIVb, Collinsella, Fusicatenibacter* and *Parabacteroides*, correlated with unhealthier eating traits and higher subjective hunger ratings. *Collinsella* was further related to higher body fat mass and *Streptococcus* to lower systolic blood pressure. The health-related bacterial genera were also more abundant in the overweight good responder controls, compared to the obese bad responder controls and RYGB-operated patients, while the inversely health-related genera showed a less clear distribution across groups, with *Parabacteroides* being significantly less abundant in good vs. bad RYGB-operated patients. Moreover, relative abundance of *Parabacteroides* as well as a composite score of all inversely correlated genera, were associated with higher eating restraint and with lower post-operative weight loss across both RYGB groups. Considering diet and SCFA-related pathways, we observed that higher dietary fiber intake in overweight adults correlated with more abundant *Clostridium XVIII*, and less abundant *Collinsella* and *Parabacteroides*, as well as with healthier eating behaviour and anthropometrics. While SCFA showed a rather mixed pattern of correlations with the different markers, *Fusicatenibacter* and *Parabacteroides* abundance correlated with higher fecal propionate and acetate, respectively, that again correlated with elevated hunger. Contrastingly, higher acetate and butyrate in serum correlated with lower fat mass, indicating a possible inverse association of acetate in feces and serum with respect to health indicators. Together, these results indicate that presumably beneficial and unfavourable microbiota genera relate to eating behaviour and weight status, and that dietary fiber intake and SCFA metabolism may modify these relationships.

### Bacterial genera

The health-related microbiota group is comprised of bacterial genera that have been described as beneficial for the host in previous literature. For example, *Alistipes* and *Blautia* were found to produce SCFA [30, 31]. Similarily, *Gemmiger, Roseburia*, and *Ruminococcus* belong to the families of Ruminococcaceae or Lachnospiraceae, which share a common role as active plant degraders [32]. These positive metabolic effects on the host could eventually contribute to improved adiposity control, as e.g. higher *Blautia* was correlated to lower body fat [33], *Roseburia* was linked to lower blood glucose and *Ruminococcus* to higher weight loss in mice after vertical sleeve gastrectomy via regulation of nuclear receptor binding of bile acids [34]. A microbial transfer study from human to mice showed obesity-promoting effects of the species *C. ramosum*, which is part of *Clostridium XVIII* [35]. However, studies on the genera *Clostridium XVIII* and *Streptococcus* in relation to health are scarce. Clostridia are known to be key commensals for gut homeostasis [36], but classification of the genus *Clostridium* remains challenging due to the high heterogeneity of the listed species [37]. Also, there are currently 50 species identified in the genus *Streptococcus* alone, rendering different functionality in these genera likely. Yet, we found that *Clostridium XVIII* abundance related to higher dietary fiber intake, and *Streptococcus* abundance to lower blood pressure. Indeed, fiber intake related to healthy eating behaviour and lower body fat mass in overweight young adults in the present analyses may point towards rather beneficial fiber-correlating *Clostridium XVIII* and *Streptococcus* genera species that underly those associations. Moreover, these results underline the potential impact of a fiber-rich diet for health indicators. Due to the exclusive occurrence of fiber in plants, fiber-rich diets are oftentimes attributed to plant-based (vegetarian or vegan) diets, and plant-based diets have been shown extensively to be beneficial for weight status, gut and overall health [38, 39].

Considering the inversely health-related group of microbiota, some genera were described to include pathogens, e.g. in *Clostridium XIVb* the species *C. piliforme*, the causative agent of Tyzzer’s disease [40] and *Parabacteroides* as an opportunistic pathogen in infectious diseases [41]. Of note, in the *Parabacteroides* genera, also beneficial species, e.g. *P. distasonis*, have been described [42]. The anaerobic *Collinsella* colonizes mucosal surfaces and has recently been reported to degrade potentially toxic food contaminants found in processed foods [43]. While this could be beneficial for the host, unhealthier eating behaviors (such as intake of processed food) and higher body weight could then likely be related to higher abundances of *Collinsella*. Likewise, studies showed that *Collinsella* linked to less dietary fiber intake, which is in line with our results in overweight adults, and higher weight loss in cross-sectional [44] and dietary intervention studies [45]. *Fusicatenibacter*, including the species *F. saccharivorans*, are strictly anaerobic sugar fermenters, again linking to unhealthier eating behaviour and obesity [46]. The genus *Clostridum IV* however has rather been reported as beneficial SCFA producers, e.g. the species *Faecalibacterium prausnitzii* (*F. prausnitzii*), which play a noticeable role in intestinal homeostasis [47]. Yet again, those genera comprise many different species and it can also be speculated that some bacteria species or genera underlying the observed correlations could have likely been taxonomically misplaced [40]. Taken together, the negatively correlating microbiota genera seem to consist on the one hand of pathogens, indicative of a rather pro-inflammatory milieu in participants with higher weight status, which is well in line with our findings showing that higher *Parabacteroides* correlated with unhealthier eating traits and poorer weight loss maintenance in RYGB patients. On the other hand, those negatively correlating genera are comprised of those bacteria that metabolize processed food and sugars, again indicative of higher weight and unhealthy eating behaviour. Future studies now need to integrate microbiota data at the species level and randomized interventional trials are required to eventually understand cause and effect of these eating behaviour-microbiota-diet interrelations.

### SCFA metabolism

We could not establish reliable links between serum and fecal concentrations of those metabolites. The overall weak relationship might be explained by rapid metabolization of SCFAs, as for example butyrate is rapidly absorbed by the gut mucosa and reaches blood circulation [48], therefore, fecal levels of butyrate may not directly relate to butyrate-producing bacteria abundance nor to serum levels of butyrate. In addition, biosamples of serum and feces were not collected in a time-locked way, therefore a time difference of hours to days might have blurred potential (anti-) correlations. Indeed, it has been shown, that fecal SCFA levels decrease throughout the day due to metabolization and that overnight-fast duration influenced these results [49].

Still, we found that higher fecal SCFA levels (i.e., acetate, butyrate and propionate) linked to higher subjective hunger ratings and also to higher cognitive restraint (i.e. propionate), whereas lower acetate and butyrate in serum correlated with higher fat mass. Statistical path analyses proposed that higher *Parabacteroides* abundance link to higher hunger through higher fecal acetate. Bearing in mind that higher fecal SCFA levels may indicate less efficient absorption in the gut, leading to lower SCFA availability in serum [50], these findings are somewhat in line with studies showing reduced appetite and less weight gain after acetate intake [1, 51]. Note however, that we did not adjust mediation statistics for multiple testing, rendering false positives likely. In addition, it has been discussed that only a minimal fraction of the colon-derived SCFA directly reaches the brain. Instead, more downstream targets of SCFA signalling might be more important for gut-brain communication, such as SCFA-induced release of GLP-1 and PYY at the gut epithelium, modulation of liver metabolism or indirect signaling via the vagus nerve [1]. Future studies could help to further disentangle the different mechanisms at play by assessing further blood-, tissue- or imaging-based biomarkers of these pathways.

## Limitations

Firstly, all analyses are based on cross-sectional data, therefore no conclusions about causal relationships can be drawn. We performed exploratory analyses centered around core hypotheses with the aim to gain more specific testable hypotheses for upcoming intervention trials. In addition, both samples are limited by size, especially with regard to the larger number of variables of interest. Due to these constraints, more elaborate statistical analyses (such as structural equation modelling) could not be performed. A major strength of this study is the inclusion of two independent samples integrating next generation sequencing and SCFA metabolomics with psychological markers in well-characterized adults at risk for future weight gain that yielded similar associations of eating behaviour with gut microbiota at the genera level.

## Conclusion

The combination of data from cross-sectional samples of overweight, obese and post-bariatric surgery individuals showed multivariate associations between specific bacterial gut genera, particularly beneficial SCFA-producing genera and presumably unfavourable pathogens or sugar-/processed-food digesting bacteria, with anthropometrics, eating traits and dietary fiber intake. While speculative concerning causality, our results propose key microbiota candidates for diet-gut-brain-behaviour interactions in humans and may help to develop novel hypotheses how to prevent and treat unhealthy food craving through microbiotal modulation of the gut-brain axis. Longitudinal and interventional studies integrating metagenomic approaches and functional pathway analysis are needed to disentangle correlation from causality and to further characterize eating behaviour-relevant microbiota genera at the species level.

## Data Availability

All data relevant to the study are included in the article or uploaded as supplementary information.

## Acknowledgments

We would like to thank all participants and staff of the study team, in particular Laura Hesse, Emmy Töws, Linda Grasser and Mathis Lammert. This work was supported by grants of the German Research Foundation, contract grant number CRC 1052 “Obesity mechanisms”, project number 209933838, subprojects A1 (AV), A5 (AH), AOBJ: 624808 (WF), WI 3342/3-1 (AVW), IFB AdiposityDiseases FKZ: 01EO1501 (AH, WF), by the Else Kröner-Fresenius Foundation (WF), by the German Federal Environmental Foundation (EM) and by the Max Planck Society.

