## Supplementary material for "Gut microbiota link dietary fiber intake and short-chain fatty acid metabolism with eating behaviour": SI (from another version of this preprint: version 1, 2021-02-20)

#### Online supplemental materials and methods

#### Methods

*Anthropometrics*

Body-mass-index (BMI) was measured as body weight (kg) divided by squared body height ($m^2$). Participants were weighed in light clothes and no shoes on in a fasted state on the same weight scale (100 g resolution, Seca GmbH, Germany) and their height was measured while standing against the wall with a fixed measuring scale (0.5 cm resolution, Seca GmbH, Germany). Waist-to-hip-ratio (WHR) was defined as ratio between waist and hip circumference, waist circumference was measured between the lower costal arch and the iliac crest, hip circumference was measured above the head of the greater trochanter (thigh bone) using a tape measure (0.1 cm resolution, Seca GmbH, Germany).

For grouping according to weight status into overweight and obesity categories, we followed the definitions of the WHO based on BMI cut-offs, except for one woman in sample 1 who showed a BMI of 31 kg/$m^2$ yet did not fall in the obesity category according to WHR-defined cut-offs (WHR = 0.78; cut-off= 0.8).

*Stool sample collection.*

Participants were asked to deliver a stool sample, which was collected 1-2 days before the testing day. A stool kit was handed out, samples were taken independently at home, where they were frozen and stored at -15 to -20°C. Samples were delivered to the institute in isolated boxes to keep the temperature until they were stored at -80°C, where they were kept until further measurement.

*Hunger ratings and health indicators.*

Hunger ratings after overnight fasting and post-prandially (10min, 40min, 65min, respectively) to a standardized meal (10% of caloric need) were taken using Visual Analogue Scales from 1 to 8 on a computer screen. Blood pressure was measured with three consecutive measurements after 10 minutes of resting in lying position with OMROM M500 HEM-7213-D (OMRON HEALTHCARE Co., Ltd., Kyoto, Japan) . The mean for each diastolic and systolic phase were calculated separately as the sum of the three measurements divided by the number of measurements. Percent body fat mass was measured using bioimpedance analysis with BIACORPUS RX 4004M (Medi Cal Healthcare GmbH, Karlsruhe, Germany) and two electrodes each at both hands and feet. Body fat was sex-standardized using z-transformation before analysis.

*Dietary Intake.*

Dietary intake was recorded using the German Food Frequency Questionnaire DEGS1 [1,2] by the Robert Koch Institute (Berlin, Germany), a tool to measure the intake of 53 single food items consumed based on self-report of frequency and quantity. Mean daily portion in grams were further used to compute fiber intake (g/day) over the last 7 days using reference food items from the German nutrition information database 'Bundeslebensmittelschlüssel' (BLS, Version 3.02) or individual sources directly from food suppliers in rare cases (e.g. for plant-based milk). Computed habitual fiber intake was normalised to 1000 kcal to adjust for individual energy intake.

*Measurement of SCFA in feces and blood.*

*Serum sample collection:* Blood was obtained in fasting state (12 ± 3h fasted) at either 7:45 am, 9:45 am or 11:45 am of each testing day using saftety-multifly-needles (21G, 200mm) and BD Vacutainer Multiple Sample Luer Adapter. Participants have been asked to fast for 12 hours before collection. Use of drugs and other confounders such as extraordinary mental or physical stress, physical activity beforehand and blood/plasma donations were recorded. Blood samples were centrifuged at 3500 revolutions per minute at 7°C for 6 minutes and serum was aliquoted within one hour of obtainment. Processed aliquots were stored at -80°C until data analysis.

*Metabolite extraction:*

Chemicals: Acetonitrile, formic acid and methanol were purchased from Sigma Aldrich (St Louis, MO, USA). D7-butyric acid was purchased from Cambridge Isotope Laboratories (Tewksbury, MA, USA). All short chain fatty acids standards (SCFAs) used for linear regression and quantitation were purchased from Sigma Aldrich (St Louis, MO, USA). All solvents for MS were of analytical grade purity. Experimental water (resistivity of 18.2 MΩ cm) was purified using a Milli-Q system (Millipore, Milford, MA, USA).

For SCFAs the method of Han et al. (2015) was modified. First, 100 mg feces were mixed with 500 µl ACN:Water (1:1, v/v) and homogenized using a TissueLyser II (30 Hz, 10 min; Retsch Qiagen). After short centrifugation (2 min, 14000 rpm) 100 µl of the supernatant were added to 500 µl ACN:Water:methanol (3:1:2, v/v/v) and the sample was vortexed for 5 min. After sonication (5 min) and centrifugation (14.000 rpm, 4°C, 5 min) 550 µl of the supernatant were transferred into a new tube and evaporated to dryness. Pellet was reconstituted in 100 µl 50% and 38 µl used for further derivatization. Next, 20 µl serum and 2 µl of standards were diluted with 18 µl and 38 µl 100% ACN, respectively. For derivatization, both specimen, serum and feces supernatant, were combined with 2 µl D7-butyric acid (2 mM) used as internal standard, 20 µl 3-nitrophenylhydrazine in 50% ACN (200 mM) and 20 µl N-(3-dimethylaminopropyl)-N'-ethylcarbodiimide hydrochloride in 50% ACN with 6% pyridine (120 mM). Incubation of the mixture was done for 30 min at 40 °C in a thermomixer (Eppendorf, Hamburg, Germany).

Prior to measurement, the resulting derivative was diluted 1:50 in 10% ACN. Of each sample 10 µl were injected into the UltiMate 3000 HPLC system (ThermoFisher Scientific™, Waltham, MA, USA) coupled online to a QTRAP® 5500 mass spectrometer (Sciex, Framingham, USA). Chromatographic separation of SCFAs was performed on an Acquity UPLC BEH C18 column (1.7µm, 2.1 x 100 mm) with H2O + 0.01 % formic acid and ACN + 0.01% formic acid as mobile phases. Constant flow rate was set to 0.35 ml and linear LC gradient was as follows: 0-2 min at 15% B, 2-17 min 15-50% B, 17-18 min 100 % B, 18-18.1 min 100-15% B, 18.1 -21 min 15 % B. Mass spectrometric measurement was performed in negative ionization mode. For identification and quantitation, a scheduled multiple reaction monitoring (MRM) method was used, with specific transitions for every SCFA. Peak areas of all samples and standards for linear regression were determined in Analyst® Software (v. 1.6.2, AB Sciex) and areas for single SCFAs were exported. Normalization and statistics were performed with in-house written R scripts.

*Bariatric surgery.*

All patients underwent standard laparoscopic Roux-en-Y gastric bypass with a 150cm alimentary und 50cm biliopancreatic limb. The indication for surgery was given interdisciplinary following the German S3- and IFSO guideline. Prior surgery, all patients underwent an hypocaloric diet, rich in protein, including commercial protein shakes, for 2 weeks.

**Supplementary Results**

*Microbiota abundance.*

In sample 1, alpha diversity varied between 89-135 (effective Shannon's index mean 110 ± 14 SD) and the phyla Actinobacteria, Bacteroidetes, Firmicutes and Proteobacteria occurred in all participants with Firmicutes being the most abundant in every participant (56-83%), followed by Bacteroidetes (12-31%) (**Figure 1A**). Fusobacteria and Verrucomicrobia occurred in some participants only. At the family level, Prevotellaceae showed considerable differences across the sample (mean 3.4% ± 4.6 SD, range 0-15%) (**Figure 1B**). At the genera level, we observed that 33-76% (mean 59.3% ± 11.2 SD) of an individual's microbiota composition stemmed from those 20 bacteria genera that were detected in at least 80% of the participants, out of which some were highly correlated (**Suppl. Figure 2-3**).

In sample 2, alpha diversity varied between 51-370 (effective Shannon's index mean 232 ± 90 SD) microbiota composition showed a higher proportion of Bacteroidetes and less Firmicutes compared to sample 1, and Prevotellaceae and Fusobacteriaceae were more abundant in obese patients after RYGB surgery (**Figure 1C-D**).

Mean alpha diversity across groups of sample 2 was similar, with large individual variability (mean 232 ± 90 SD). The 20 highly prevalent genera detected in sample 1 showed a relatively high variation and accounted for 6-85% of bacteria abundance across participants (overweight: mean 64.7% ± 19.5 SD, obese: mean 54.1% ± 20.9 SD, good RYGB responders 26.4% ± 14.1 SD, bad RYGB responders 35.4% ± 18.7 SD). Note that Clostridium *XVIII* were found in 40 % of the participants only, *Fusicatenibacter* in 65% only.

*Mediation analyses.*

Simple mediation path analyses gave mixed results: the effects of fiber on eating behaviour mediated by sumscores of microbiotal genera, did not show significant associations (for total effects all $0.08 < |Z| > 1.5$, all p-uncorr < 0.94). For direct effects of eating traits on genera sumscore mediated by fiber intake, cognitive restraint and disinhibition significantly affected negative genera sumscore (all $Z > 3.5$, $p < 0.001$), and disinhibited eating and hunger trait significantly increased *Parabacteroides* abundance (all $Z > 2.7$, $p < 0.01$). Moreover, direct effects of hunger on positive sumscore were present, showing that higher hunger affected lower positive genera abundance (all $Z > -2.2$, $p < 0.05$).

### Supplementary Figures

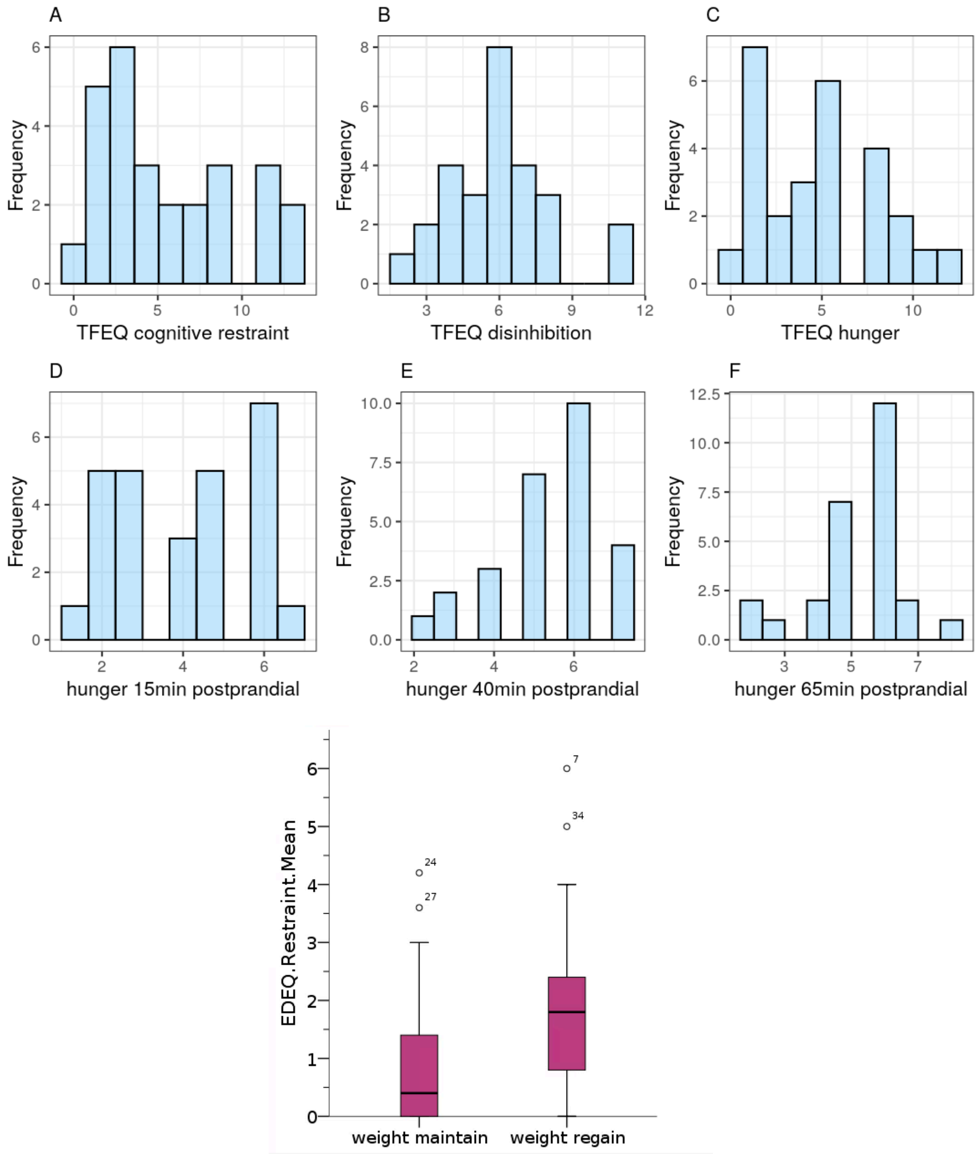

Suppl. Figure 1: Distribution of eating behaviour traits in overweight adults (sample 1) and between good and bad RYGB responders (sample 2).

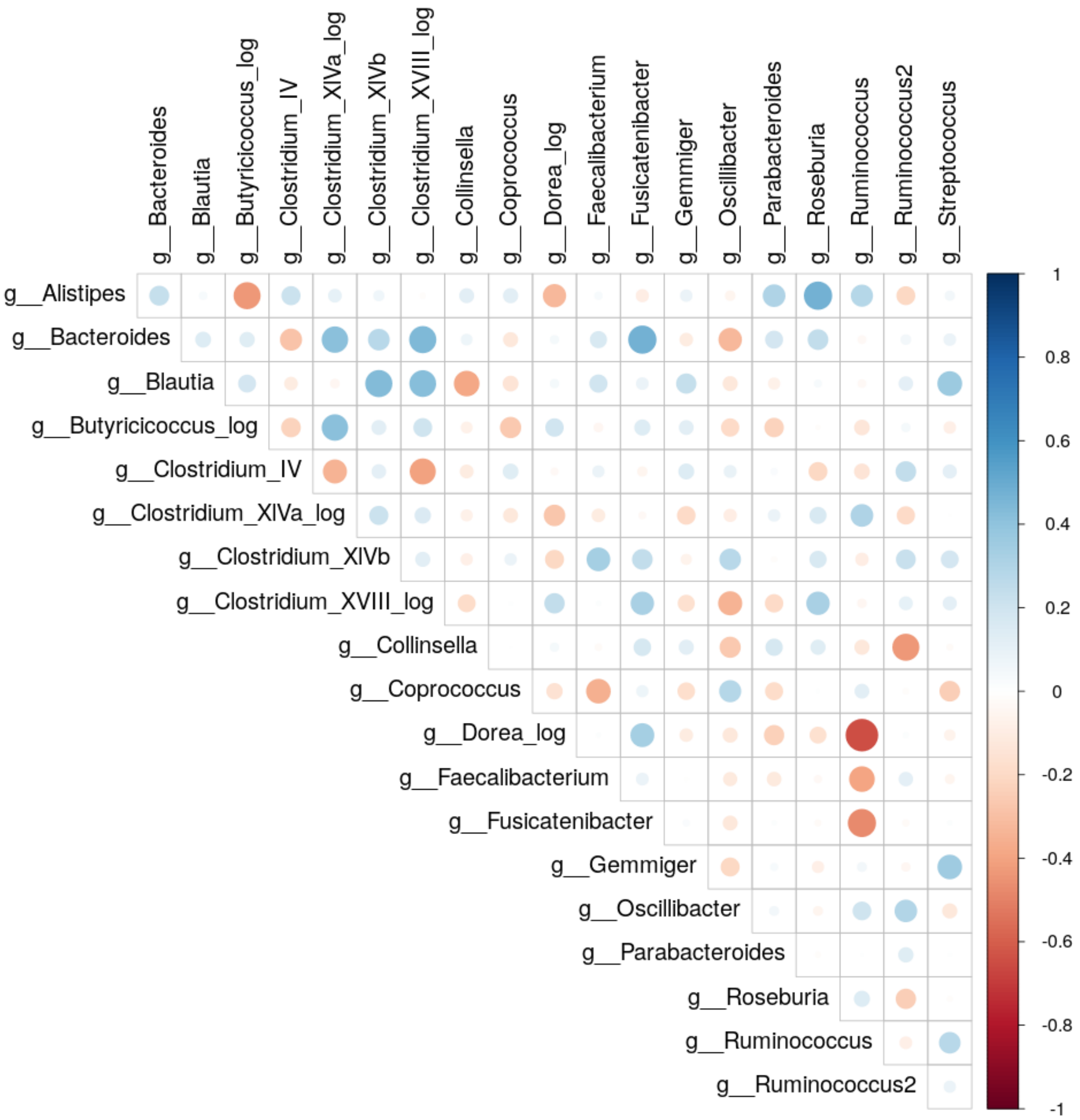

Suppl. Figure 2: Collinearities between the 20 genera that were detected in at least 80% of participants of sample 1. Of those, 7 were health-related (Alistipes, Blautia, Clostridium XVIII, Gemmiger, Roseburia, Ruminococcus, Streptococcus) and 5 inversely health-related (Clostridium IV, Clostridium XIVb, Collinsella, Fusicatenibacter, Parabacteroides) bacterial genera, according to correlation analyses in sample 1 (Pearson's r, p-uncorrected, n = 27, sample 1).

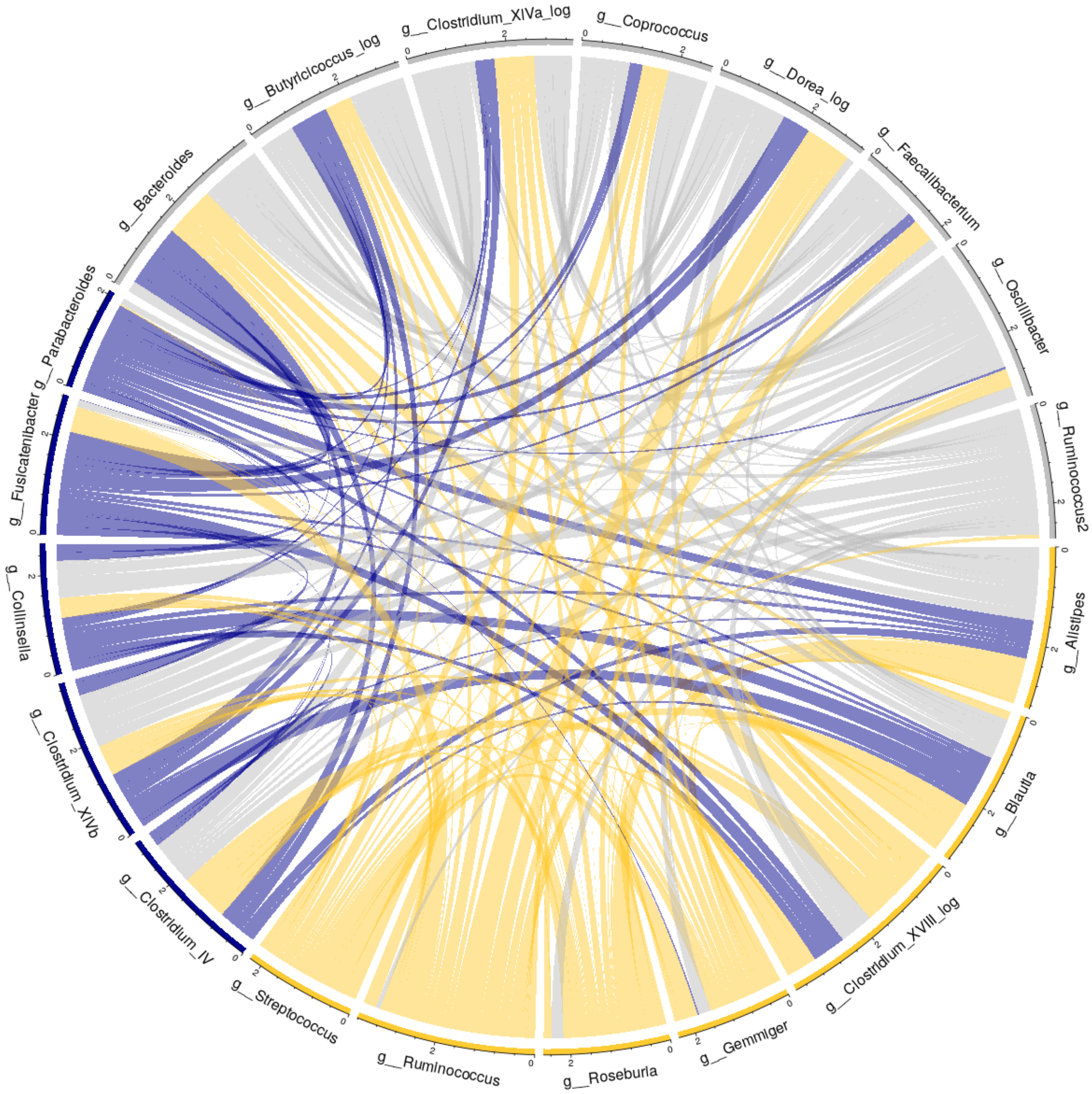

Suppl. Figure 3: Collinearities between 20 genera that were detected in at least 80% of participants of sample 1 (n=27). Chord Diagram (Pearson's r), blue: inversely health-related genera; yellow: health-related genera; grey: eating behaviour-non-related genera.

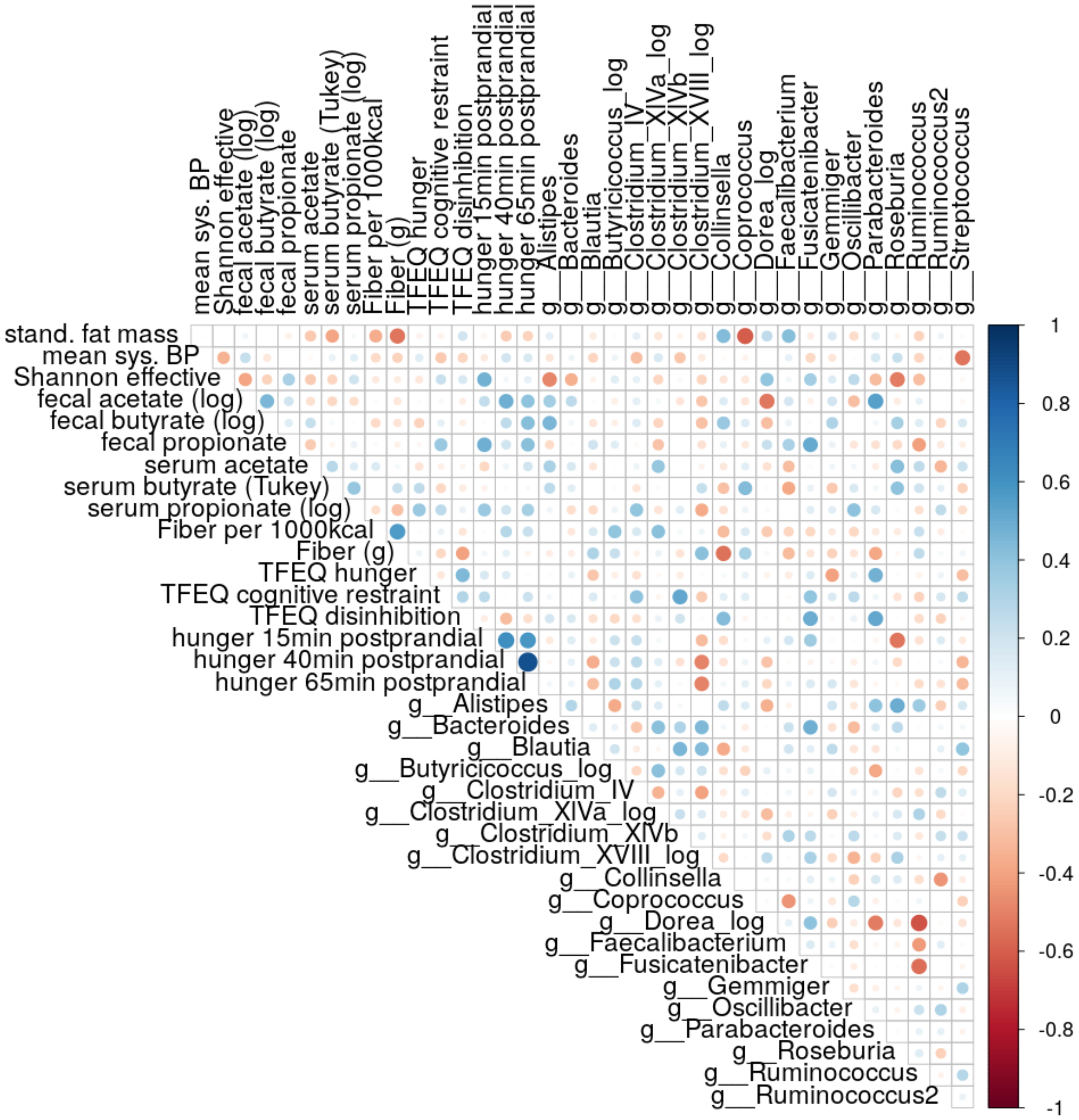

Suppl. Figure 4: Corrplot for all variables of interest (subjects n = 27, sample 1), Pearson's r and p-uncorrected.

| | fecal butyrate (log) | fecal propionate | serum acetate | serum butyrate (Tukey) | serum propionate (log) | g__Alistipes | g__Blautia | g__Clostridium_IV | g__Clostridium_XIVb | g__Clostridium_XVIII (log) | g__Collinsella | g__Fusicatenibacter | g__Gemmiger | g__Parabacteroides | g__Roseburia | g__Ruminococcus | g__Streptococcus |
|---|---|---|---|---|---|---|---|---|---|---|---|---|---|---|---|---|---|
| fecal acetate (log) | 0.45 | 0.19 | -0.17 | -0.2 | -0.17 | 0.34 | 0.02 | 0.12 | -0.14 | -0.28 | 0.15 | -0.08 | 0.2 | 0.55 | -0.1 | 0.1 | -0.04 |
| fecal butyrate (log) | | 0.14 | 0.23 | 0.04 | -0.01 | 0.46 | -0.14 | 0.15 | 0 | -0.3 | 0.38 | -0.02 | 0.31 | 0.05 | 0.34 | 0.14 | 0.17 |
| fecal propionate | | | -0.26 | -0.06 | 0.09 | -0.18 | 0.2 | -0.04 | 0 | -0.16 | 0.1 | 0.5 | 0.09 | -0.16 | -0.19 | -0.41 | -0.09 |
| serum acetate | | | | 0.28 | 0.14 | 0.33 | -0.13 | 0.07 | -0.01 | -0.07 | -0.12 | -0.02 | 0.06 | -0.09 | 0.42 | 0.26 | 0.22 |
| serum butyrate (Tukey) | | | | | 0.38 | 0.26 | 0.01 | -0.14 | -0.01 | 0.23 | -0.29 | 0.09 | -0.26 | 0.06 | 0.4 | 0.21 | -0.23 |
| serum propionate (log) | | | | | | 0.06 | -0.18 | 0.39 | 0.14 | -0.38 | -0.16 | 0.13 | 0.12 | 0.18 | 0.02 | -0.16 | -0.17 |

Suppl. Figure 5: Correlations between fecal and serum levels of SCFA (acetate, butyrate and propionate) and bacterial genera of interest (Pearson's r, n = 27, sample 1) (blue: inversely health-related genera; yellow: health-related genera).

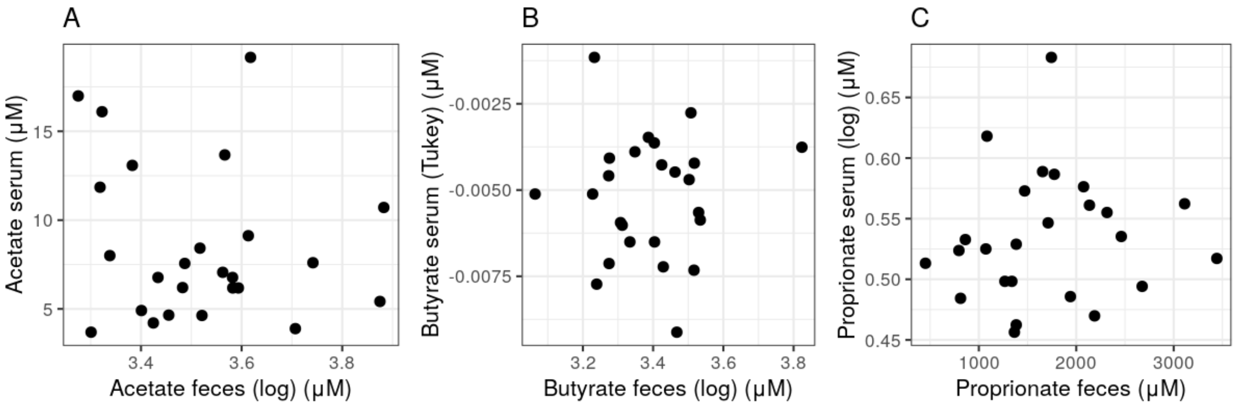

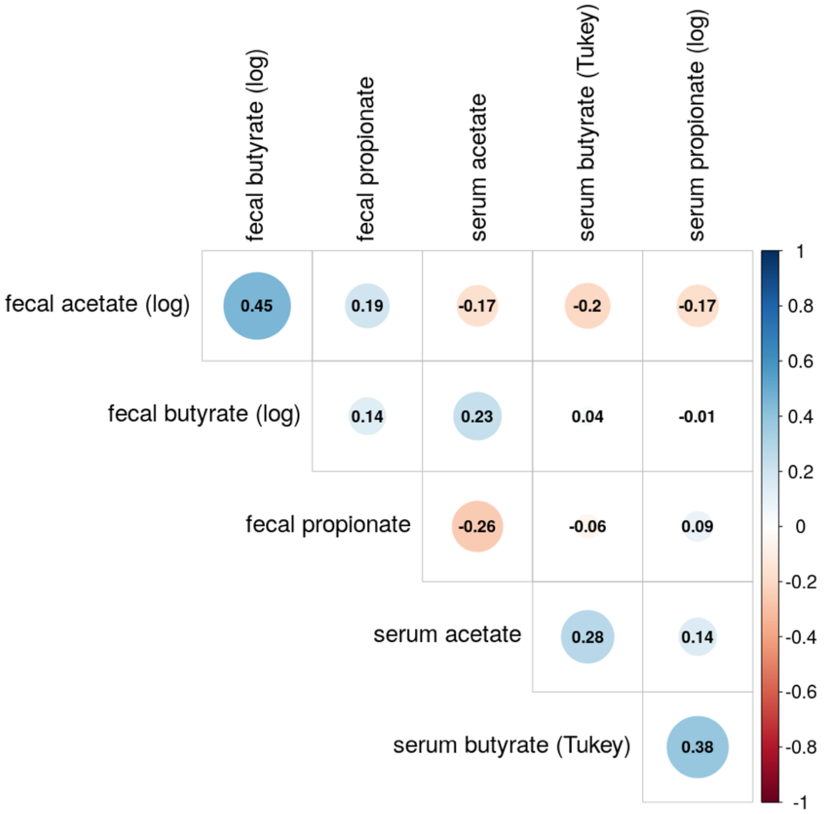

Suppl. Figure 6: Associations between serum and feces levels of SCFA for acetate, butyrate and propionate (Pearson, n = 27, sample 1).

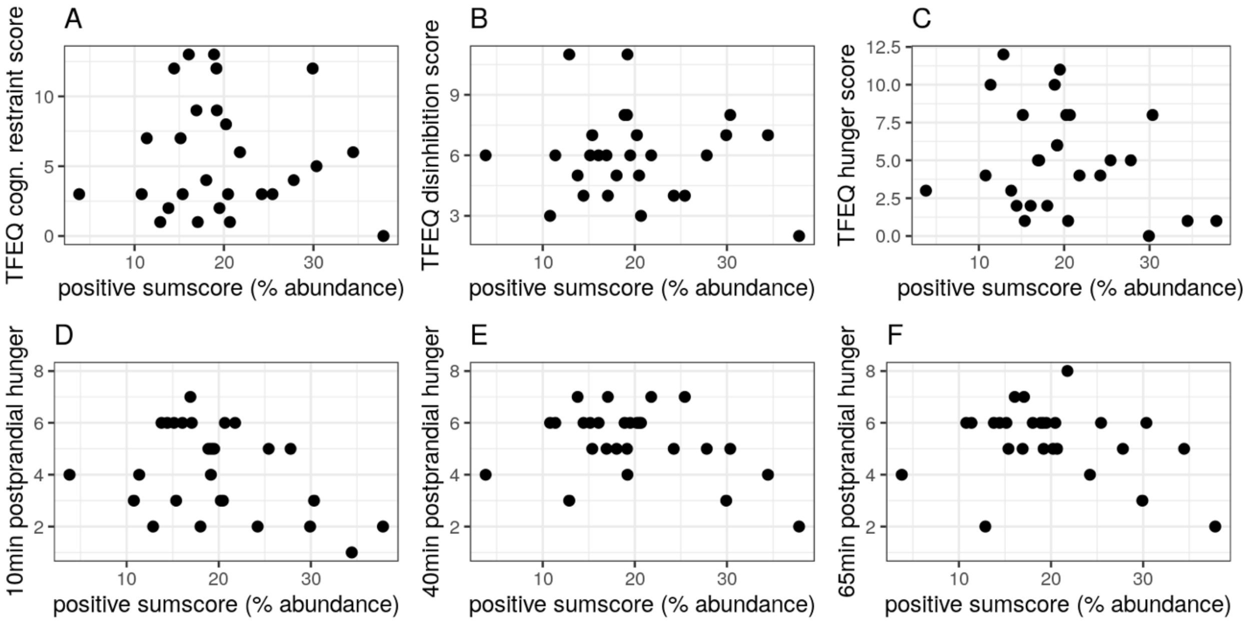

Suppl. Figure 7: Associations microbial genera sum score for health-related genera with eating behaviour outcomes (Pearson's r, p-uncorrected, n = 27, sample 1).

Suppl. Table 1: Concentrations of feces and serum SCFA levels in sample 1.

| | *Sample 1* | | | | |
|---|---|---|---|---|---|
| | *mean* | *SD* | *minimum* | *maximum* | *paired t-test between feces and serum* |
| *Feces Acetate [µmol]* | 3603 | 1534 | 1884 | 7630 | t(24) = 11.7, p < 0.001 |
| *Feces Butyrate [µmol]* | 2687 | 1051 | 1158 | 6663 | t(24) = 12.4, p < 0.001 |
| *Feces Propionate [µmol]* | 1835 | 854 | 452 | 3511 | t(24) = 11.6, p < 0.001 |
| *Serum Acetate [µmol]* | 8.5 | 4.3 | 3.7 | 19.2 | |
| *Serum Butyrate [µmol]* | 2.5 | 0.4 | 2.0 | 3.9 | |
| *Serum Propionate [µmol]* | 2.5 | 0.4 | 1.9 | 3.8 | |

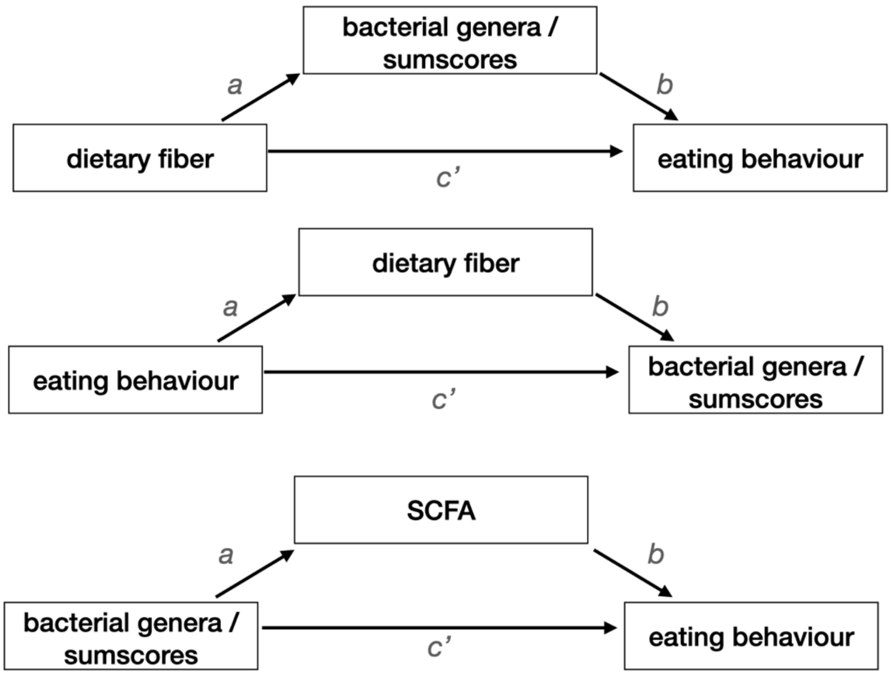

Suppl. Figure 8: Proposed mediation models.

Suppl. Table 2: Mediation effects of fiber intake on eating behaviour (via bacterial genera) in sample 1.

| | TFEQ hunger | | TFEQ cognitive restraint | | TFEQ disinhibition | | Hunger 15 min postprandial | | Hunger 40 min postprandial | | Hunger 65 min postprandial | |
|---|---|---|---|---|---|---|---|---|---|---|---|---|
| **Model 1 (via positive genera sumscore)** | *ß* | *p* | *ß* | *p* | *ß* | *p* | *ß* | *p* | *ß* | *p* | *ß* | *p* |
| Mediation effect *a*b* (fiber on behaviour via genera) | -0.02 | 0.74 | -0.00 | 0.86 | -0.01 | 0.79 | -0.02 | 0.74 | -0.03 | 0.74 | -0.02 | 0.74 |
| Direct effect *c′* | 0.00 | 0.99 | 0.08 | 0.68 | -0.12 | 0.53 | 0.06 | 0.77 | 0.31 | 0.08 | 0.25 | 0.19 |
| Total effect *c + a*b* (fiber on behaviour) | -0.16 | 0.94 | 0.08 | 0.69 | -0.13 | 0.51 | 0.03 | 0.89 | 0.29 | 0.13 | 0.23 | 0.24 |
| **Model 2 (via negative genera sumscore)** | | | | | | | | | | | | |
| Mediation effect *a*b* (fiber on behaviour via genera) | -0.04 | 0.45 | -0.11 | 0.34 | -0.12 | 0.33 | -0.03 | 0.54 | -0.01 | 0.74 | -0.05 | 0.44 |
| Direct effect *c′* | 0.03 | 0.89 | 0.19 | 0.24 | -0.01 | 0.95 | 0.06 | 0.77 | 0.30 | 0.12 | 0.27 | 0.16 |
| Total effect *c + a*b* (fiber on behaviour) | -0.02 | 0.94 | 0.08 | 0.69 | -0.13 | 0.51 | 0.03 | 0.89 | 0.29 | 0.13 | 0.23 | 0.24 |

Suppl. Table 3: Mediation effects of eating behaviour on bacterial genera (via fiber intake) in sample 1.

| | **positive genera sumscore** | | **negative genera sumscore** | | **Parabacteroides abundance** | |
|---|---|---|---|---|---|---|
| **Model 1** **(TFEQ hunger)** | *ß* | *p* | *ß* | *p* | *ß* | *p* |
| Mediation effect *a*b* (TFEQ on genera via fiber) | 0.00 | 0.92 | 0.00 | 0.94 | 0.00 | 0.94 |
| Direct effect *c′* | 0.23 | 0.25 | 0.22 | 0.25 | 0.47 | **0.007** |
| Total effect *c + a*b* (TFEQ hunger on genera) | 0.23 | 0.24 | 0.22 | 0.25 | 0.47 | **0.07** |
| **Model 2** **(TFEQ cognitive restraint)** | | | | | | |
| Mediation effect *a*b* (TFEQ on genera via fiber) | 0.01 | 0.79 | -0.02 | 0.70 | -0.01 | 0.74 |
| Direct effect *c′* | -0.04 | 0.84 | 0.57 | **<0.001** | 0.07 | 0.71 |
| Total effect *c + a*b* (TFEQ on genera) | -0.04 | 0.86 | 0.55 | **<0.001** | 0.06 | 0.75 |
| **Model 3** **(TFEQ disinhibition)** | | | | | | |
| Mediation effect *a*b* (TFEQ on genera via fiber) | -0.01 | 0.80 | 0.02 | 0.62 | 0.00 | 0.80 |
| Direct effect *c′* | -0.10 | 0.65 | 0.62 | **<0.001** | 0.51 | **0.003** |
| Total effect *c + a*b* (TFEQ on genera) | -0.10 | 0.62 | 0.63 | **<0.001** | 0.51 | **0.003** |
| **Model 4** **(hunger 10 min postprandial)** | | | | | | |
| Mediation effect *a*b* (hunger on genera via fiber) | 0.00 | 0.89 | -0.01 | 0.89 | 0.00 | 0.89 |
| Direct effect *c′* | -0.40 | **0.03** | 0.16 | 0.42 | -0.05 | 0.79 |
| Total effect *c + a*b* (hunger on genera) | -0.39 | **0.03** | 0.15 | 0.44 | -0.06 | 0.78 |
| **Model 5** **(hunger 40 min postprandial)** | | | | | | |
| Mediation effect *a*b* (hunger on genera via fiber) | 0.05 | 0.41 | -0.06 | 0.39 | -0.04 | 0.53 |

| | | | | | | |
|---|---|---|---|---|---|---|
| Direct effect *c'* | -0.42 | **0.03** | 0.07 | 0.72 | 0.10 | 0.64 |
| Total effect *c + a*b* (hunger on genera) | -0.37 | **0.05** | 0.01 | 0.95 | 0.06 | 0.78 |
| **Model 6 (hunger 65 min postprandial)** | | | | | | |
| Mediation effect *a*b* (hunger on genera via fiber) | 0.03 | 0.56 | -0.06 | 0.39 | -0.03 | 0.62 |
| Direct effect *c'* | -0.29 | 0.15 | 0.24 | 0.22 | -0.02 | 0.92 |
| Total effect *c + a*b* (hunger on genera) | -0.26 | 0.19 | 0.18 | 0.36 | -0.05 | 0.82 |

Suppl. Table 4: Mediation effects of bacterial genera on eating behaviour (via SCFA) in sample 1.

| | **positive genera sumscore** | | **negative genera sumscore** | | **Parabacteroides abundance** | |
|---|---|---|---|---|---|---|
| **Model 1 (dep: TFEQ cognitive constraint; med: fecal Propionate)** | *ß* | *p* | *ß* | *p* | *ß or 95% CI* | *p* |
| Mediation effect *a*b* (genera on TFEQ via SCFA) | -0.06 | 0.44 | 0.05 | 0.44 | -0.06 | 0.46 |
| Direct effect *c'* | 0.03 | 0.88 | 0.50 | **0.002** | 0.13 | 0.50 |
| Total effect *c + a*b* (genera on TFEQ) | -0.04 | 0.86 | 0.55 | **<0.001** | 0.06 | 0.75 |
| **Model 2 (dep: postprandial hunger; med: fecal Acetate)** | | | | | | |
| Mediation effect *a*b* (genera on hunger via SCFA) | 0.04 | 0.71 | 0.17 | 0.15 | 0.36 **[0.05 0.66]** | **0.02** |
| Direct effect *c'* | -0.40 | **0.01** | -0.15 | 0.39 | -0.30 | 0.13 |
| Total effect *c + a*b* (genera on hunger) | -0.37 | **0.05** | 0.01 | 0.95 | 0.06 | 0.78 |
| **Model 3 (dep: postprandial hunger; med: fecal Butyrate)** | | | | | | |
| Mediation effect *a*b* (genera on hunger via SCFA) | 0.15 | 0.19 | 0.09 | 0.30 | 0.02 | 0.79 |
| Direct effect *c'* | -0.40 | **0.02** | 0.09 | 0.63 | -0.07 | 0.70 |

| | | | | | | |
|---|---|---|---|---|---|---|
| Total effect *c + a*b* (genera on hunger) | -0.26 | 0.19 | 0.18 | 0.36 | -0.05 | 0.82 |
| **Model 4 (dep: postprandial hunger; med: fecal Propionate)** | | | | | | |
| Mediation effect *a*b* (genera on hunger via SCFA) | -0.07 | 0.42 | 0.08 | 0.42 | -0.08 | 0.45 |
| Direct effect *c′* | -0.32 | 0.052 | 0.07 | 0.68 | 0.20 | 0.91 |
| Total effect *c + a*b* (genera on hunger) | -0.39 | **0.03** | 0.15 | 0.44 | -0.06 | 0.78 |
| **Model 5 (dep: postprandial hunger; med: serum Propionate)** | | | | | | |
| Mediation effect *a*b* (genera on hunger via SCFA) | -0.05 | 0.50 | 0.10 | 0.25 | 0.07 | 0.41 |
| Direct effect *c′* | -0.35 | **0.047** | 0.05 | 0.79 | -0.13 | 0.50 |
| Total effect *c + a*b* (genera on hunger) | -0.39 | **0.03** | 0.15 | 0.44 | -0.06 | 0.78 |

Suppl. Table 5: Correlation matrix for all variables of interest (Pearson correlation, p-FDR corrected < 0.05, sorted alphabetically).

| Variable 1 | Variable 2 | cor | p | p_FDR |
|---|---|---|---|---|
| **Acetate_log** | **Butyrate_log** | 0.598859207013339 | 9.01E-05 | 0.001395892765718 |
| **Acetate_log** | **g__Alistipes** | 0.440661171890756 | 0.006339443277375 | 0.036713645415058 |
| **Acetate_log** | **g__Clostridium_XVIII_log** | -0.445473726657783 | 0.005726622472821 | 0.033751597937157 |
| **Acetate_log** | **g__Dorea_log** | -0.623880443719398 | 3.70E-05 | 0.000770182578778 |
| **Acetate_log** | **g__Parabacteroides** | 0.561751017123597 | 0.00029713238755 | 0.003664632779784 |
| **Acetate_log** | **hunger_post_wanting** | 0.56353185584319 | 0.000281499686374 | 0.003537335681609 |
| **Acetate_log** | **hunger_pre_memory** | 0.51895071561856 | 0.00099860519866 | 0.009501015175819 |
| **Acetate_serum** | **Butyrate_serum_tukey** | 0.49340489913932 | 0.001915064534996 | 0.014830616050088 |
| **Acetate_serum** | **g__Alistipes** | 0.541644422396596 | 0.000535800022944 | 0.005664171671123 |
| **Acetate_serum** | **g__Clostridium_XlVa_log** | 0.510355010058988 | 0.001250254688114 | 0.011102261630452 |
| **Acetate_serum** | **g__Faecalibacterium** | -0.560776108680864 | 0.000306015517875 | 0.003705569725542 |
| **Acetate_serum** | **g__Roseburia** | 0.636656349838231 | 2.27E-05 | 0.000605900930198 |
| **Acetate_serum** | **g__Ruminococcus** | 0.578197633425228 | 0.000178239985857 | 0.002473079803759 |
| **Acetate_serum** | **g__Ruminococcus2** | -0.456201709797121 | 0.004541513167263 | 0.029083151628822 |
| **Blutdruck..mmHg..mean_sys** | **g__Streptococcus** | -0.70274328205789 | 1.24E-06 | 5.74E-05 |
| **Blutdruck..mmHg..mean_sys** | **TFEQ_cognitive_restraint_sum** | -0.468085681931573 | 0.003482361204211 | 0.023909820226852 |
| **Butyrate_log** | **g__Alistipes** | 0.646003039383035 | 1.57E-05 | 0.000455206953737 |
| **Butyrate_log** | **g__Clostridium_XVIII_log** | -0.464528186867958 | 0.003774178903472 | 0.024643168134438 |
| **Butyrate_log** | **g__Collinsella** | 0.497254139423969 | 0.001741626952091 | 0.014145409147468 |
| **Butyrate_log** | **g__Dorea_log** | -0.487558544436211 | 0.002207493604466 | 0.015980334136678 |

| | | | | |
|---|---|---|---|---|
| **Butyrate_log** | **g__Gemmiger** | 0.522105729164989 | 0.000918167668988 | 0.009126860709646 |
| **Butyrate_log** | **hunger_pre_memory** | 0.451291356106825 | 0.005054587842143 | 0.030883995439151 |
| **Butyrate_serum_tukey** | **Fibre_g** | 0.431880293624311 | 0.007603689030378 | 0.041851709869682 |
| **Butyrate_serum_tukey** | **g__Coprococcus** | 0.66477034458707 | 7.20E-06 | 0.000281256296857 |
| **Butyrate_serum_tukey** | **g__Faecalibacterium** | -0.640877067540008 | 1.93E-05 | 0.000535003627883 |
| **Butyrate_serum_tukey** | **g__Gemmiger** | -0.482852750954815 | 0.002470574357283 | 0.017692500236025 |
| **Butyrate_serum_tukey** | **g__Roseburia** | 0.516013197878123 | 0.001079038833035 | 0.010040557420176 |
| **Fibre_g** | **g__Clostridium_XVIII_log** | 0.48823182203372 | 0.002171935841722 | 0.015895706270186 |
| **Fibre_g** | **g__Collinsella** | -0.729283828433059 | 3.06E-07 | 2.44E-05 |
| **Fibre_g** | **g__Coprococcus** | 0.453372024462275 | 0.004831372288745 | 0.030355603248152 |
| **Fibre_g** | **g__Faecalibacterium** | -0.497830088743999 | 0.001716900726402 | 0.014116739305972 |
| **Fibre_g** | **g__Parabacteroides** | -0.510853642145674 | 0.001234257868588 | 0.011102261630452 |
| **Fibre_g** | **TFEQ_disinhibition_sum** | -0.583255134232137 | 0.000151479520253 | 0.002241896899745 |
| **Fibre_per_1000kcal** | **Fibre_g** | 0.760556768527585 | 4.70E-08 | 7.86E-06 |
| **Fibre_per_1000kcal** | **g__Butyricicoccus_log** | 0.492303070121502 | 0.001967421677563 | 0.01488980496883 |
| **Fibre_per_1000kcal** | **g__Clostridium_XIVa_log** | 0.543042767415546 | 0.000514887922237 | 0.005554794801279 |
| **Fibre_per_1000kcal** | **g__Collinsella** | -0.542891598987598 | 0.000517113029548 | 0.005554794801279 |
| **Fibre_per_1000kcal** | **g__Faecalibacterium** | -0.431218154651246 | 0.007707217425172 | 0.042041454192077 |
| **Fibre_per_1000kcal** | **TFEQ_disinhibition_sum** | -0.437433663456543 | 0.006781328876634 | 0.038601410528532 |
| **FM_stand** | **Acetate_serum** | -0.433763770640638 | 0.007315690547339 | 0.040943276508637 |
| **FM_stand** | **Butyrate_serum_tukey** | -0.633315746191267 | 2.59E-05 | 0.000638308524086 |
| **FM_stand** | **Fibre_g** | -0.749535572974836 | 9.39E-08 | 8.93E-06 |
| **FM_stand** | **Fibre_per_1000kcal** | -0.633319850171976 | 2.59E-05 | 0.000638308524086 |
| **FM_stand** | **g__Collinsella** | 0.606313033588064 | 6.97E-05 | 0.001189444648582 |
| **FM_stand** | **g__Coprococcus** | -0.760249969277725 | 4.80E-08 | 7.86E-06 |
| **FM_stand** | **g__Faecalibacterium** | 0.663517615374537 | 7.60E-06 | 0.000281256296857 |
| **g__Alistipes** | **g__Butyricicoccus_log** | -0.581752860754415 | 0.000159023570996 | 0.002302384745288 |
| **g__Alistipes** | **g__Dorea_log** | -0.623847758747502 | 3.70E-05 | 0.000770182578778 |
| **g__Alistipes** | **g__Parabacteroides** | 0.558082359055829 | 0.000331808681737 | 0.003946153250663 |
| **g__Alistipes** | **g__Roseburia** | 0.646395930617989 | 1.55E-05 | 0.000455206953737 |
| **g__Alistipes** | **g__Ruminococcus** | 0.550191686692032 | 0.000418918341904 | 0.004894730100138 |
| **g__Bacteroides** | **g__Clostridium_IV** | -0.505566155517767 | 0.001413453741196 | 0.012068720405599 |
| **g__Bacteroides** | **g__Clostridium_XIVa_log** | 0.515786925319385 | 0.001085465667046 | 0.010040557420176 |
| **g__Bacteroides** | **g__Clostridium_XVIII_log** | 0.524779247271871 | 0.000854553881407 | 0.008623225530562 |
| **g__Bacteroides** | **g__Oscillibacter** | -0.499015436139158 | 0.00166698132597 | 0.013877619538696 |
| **g__Bacteroides** | **g__Roseburia** | 0.451369893036369 | 0.005046003279644 | 0.030883995439151 |
| **g__Blautia** | **g__Clostridium_XIVb** | 0.54713375494089 | 0.000457818976767 | 0.005257024802187 |
| **g__Blautia** | **g__Clostridium_XVIII_log** | 0.62627106121299 | 3.38E-05 | 0.000770182578778 |
| **g__Blautia** | **g__Collinsella** | -0.427834428668583 | 0.008255338354299 | 0.043635359872725 |
| **g__Blautia** | **g__Streptococcus** | 0.601809058772353 | 8.14E-05 | 0.00132306457214 |
| **g__Butyricicoccus_log** | **g__Clostridium_XIVa_log** | 0.425513958360277 | 0.008650209587631 | 0.045322108669325 |
| **g__Butyricicoccus_log** | **g__Parabacteroides** | -0.599195058159242 | 8.91E-05 | 0.001395892765718 |
| **g__Clostridium_IV** | **g__Clostridium_XIVa_log** | -0.492610799131712 | 0.001952673981899 | 0.01488980496883 |
| **g__Clostridium_IV** | **g__Clostridium_XVIII_log** | -0.64725190136129 | 1.49E-05 | 0.000455206953737 |
| **g__Clostridium_IV** | **g__Roseburia** | -0.444087908455195 | 0.005897617800715 | 0.034454503993649 |
| **g__Clostridium_XIVa_log** | **g__Ruminococcus** | 0.46588052083691 | 0.003660837928401 | 0.02438118060315 |

| | | | | |
|---|---|---|---|---|
| **g__Collinsella** | **g__Ruminococcus2** | -0.545861354874867 | 0.000474932493709 | 0.005361102386618 |
| **g__Coprococcus** | **g__Faecalibacterium** | -0.716343238894069 | 6.17E-07 | 4.08E-05 |
| **g__Dorea_log** | **g__Fusicatenibacter** | 0.612455444495711 | 5.61E-05 | 0.001008911128717 |
| **g__Dorea_log** | **g__Parabacteroides** | -0.579857988268415 | 0.000169020172869 | 0.00239505181129 |
| **g__Dorea_log** | **g__Ruminococcus** | -0.767505351526908 | 2.98E-08 | 7.86E-06 |
| **g__Faecalibacterium** | **g__Ruminococcus** | -0.622609710143205 | 3.87E-05 | 0.000781930777352 |
| **g__Fusicatenibacter** | **g__Ruminococcus** | -0.72797300207441 | 3.29E-07 | 2.44E-05 |
| **g__Gemmiger** | **g__Streptococcus** | 0.490273545996011 | 0.002067156159659 | 0.015468831486885 |
| **g__Oscillibacter** | **g__Ruminococcus2** | 0.429936802170668 | 0.007910995613914 | 0.042489702248923 |
| **hunger_post_wanting** | **g__Blautia** | -0.488241691107645 | 0.00217141838119 | 0.015895706270186 |
| **hunger_post_wanting** | **g__Clostridium_XVIII_log** | -0.619022772110668 | 4.42E-05 | 0.000865949116242 |
| **hunger_post_wanting** | **g__Streptococcus** | -0.481002321968606 | 0.00258130186202 | 0.01828879829899 |
| **hunger_post_wanting** | **hunger_pre_memory** | 0.926195261615634 | 2.22E-16 | 1.48E-13 |
| **hunger_pre_memory** | **g__Blautia** | -0.465353859128006 | 0.003704621454586 | 0.024428493948059 |
| **hunger_pre_memory** | **g__Clostridium_XVIII_log** | -0.655845603875821 | 1.05E-05 | 0.000368416470372 |
| **hunger_pre_memory** | **g__Streptococcus** | -0.4480000409974 | 0.005425881346766 | 0.032555288080595 |
| **hunger_pre_wanting** | **g__Alistipes** | -0.46280451701447 | 0.00392305196277 | 0.025366530167034 |
| **hunger_pre_wanting** | **g__Clostridium_IV** | 0.422336664071304 | 0.009216985609871 | 0.047585367567243 |
| **hunger_pre_wanting** | **g__Clostridium_XVIII_log** | -0.495852361320298 | 0.001803119179798 | 0.014199197996351 |
| **hunger_pre_wanting** | **g__Roseburia** | -0.756992801739824 | 5.90E-08 | 7.86E-06 |
| **hunger_pre_wanting** | **g__Ruminococcus** | -0.425167346486677 | 0.00871055541993 | 0.045322108669325 |
| **hunger_pre_wanting** | **hunger_post_wanting** | 0.703559991799895 | 1.19E-06 | 5.74E-05 |
| **hunger_pre_wanting** | **hunger_pre_memory** | 0.704780682681857 | 1.12E-06 | 5.74E-05 |
| **Propionate** | **Acetate_serum** | -0.500992384803234 | 0.001586560168223 | 0.013375304709323 |
| **Propionate** | **g__Alistipes** | -0.438892713068135 | 0.006578394602837 | 0.037769058668014 |
| **Propionate** | **g__Clostridium_XIVa_log** | -0.430855610659581 | 0.007764412711149 | 0.042041454192077 |
| **Propionate** | **g__Dorea_log** | 0.429481154229603 | 0.007984562382702 | 0.042541748375038 |
| **Propionate** | **g__Faecalibacterium** | 0.51393809446456 | 0.001139258666842 | 0.010393784549544 |
| **Propionate** | **g__Fusicatenibacter** | 0.6240426731233 | 3.67E-05 | 0.000770182578778 |
| **Propionate** | **g__Roseburia** | -0.506017325395259 | 0.001397315727119 | 0.012068720405599 |
| **Propionate** | **g__Ruminococcus** | -0.698408329392037 | 1.54E-06 | 6.40E-05 |
| **Propionate** | **hunger_pre_memory** | 0.506333126884965 | 0.001386116565458 | 0.012068720405599 |
| **Propionate** | **hunger_pre_wanting** | 0.7019207845956 | 1.29E-06 | 5.74E-05 |
| **Propionate** | **TFEQ_cognitive_restraint_sum** | 0.495648703048968 | 0.001812209954489 | 0.014199197996351 |
| **Propionate_serum_log** | **g__Blautia** | -0.448547043699575 | 0.005362590468966 | 0.032468047748467 |
| **Propionate_serum_log** | **g__Clostridium_IV** | 0.51902096277824 | 0.000996748495826 | 0.009501015175819 |
| **Propionate_serum_log** | **g__Clostridium_XVIII_log** | -0.572391316765886 | 0.000214142406679 | 0.002750091324581 |
| **Propionate_serum_log** | **g__Oscillibacter** | 0.615257076711025 | 5.07E-05 | 0.000937689476897 |
| **Propionate_serum_log** | **hunger_pre_wanting** | 0.452684889881284 | 0.004904126435014 | 0.030524749586164 |
| **shannon.effective** | **Acetate_serum** | -0.468374690308861 | 0.003459540580359 | 0.023909820226852 |
| **shannon.effective** | **g__Alistipes** | -0.71469430140524 | 6.73E-07 | 4.08E-05 |
| **shannon.effective** | **g__Bacteroides** | -0.466630508548113 | 0.003599264930892 | 0.02438118060315 |
| **shannon.effective** | **g__Clostridium_XIVa_log** | -0.445587811307233 | 0.005712738021074 | 0.033751597937157 |
| **shannon.effective** | **g__Dorea_log** | 0.603443040301911 | 7.70E-05 | 0.001281651388953 |
| **shannon.effective** | **g__Fusicatenibacter** | 0.496749226577845 | 0.00176356036501 | 0.014150978350561 |
| **shannon.effective** | **g__Parabacteroides** | -0.432627688164367 | 0.007488264531307 | 0.041559868148753 |

| | | | | |
|---|---|---|---|---|
| **shannon.effective** | **g__Roseburia** | -0.752114010608859 | 8.01E-08 | 8.89E-06 |
| **shannon.effective** | **g__Ruminococcus** | -0.583577189432241 | 0.00014990476626 | 0.002241896899745 |
| **shannon.effective** | **hunger_pre_wanting** | 0.648421377301863 | 1.43E-05 | 0.000455206953737 |
| **shannon.effective** | **Propionate** | 0.611054313658948 | 5.89E-05 | 0.001032682742647 |
| **shannon.effective** | **TFEQ_cognitive_restraint_sum** | 0.527681444686461 | 0.000789948927496 | 0.008093938241727 |
| **TFEQ_cognitive_restraint_sum** | **g__Clostridium_IV** | 0.573729336088943 | 0.000205337930194 | 0.002735101230182 |
| **TFEQ_cognitive_restraint_sum** | **g__Clostridium_XlVb** | 0.57414425484823 | 0.000202674450445 | 0.002735101230182 |
| **TFEQ_cognitive_restraint_sum** | **g__Fusicatenibacter** | 0.465897394870422 | 0.003659442589486 | 0.02438118060315 |
| **TFEQ_cognitive_restraint_sum** | **g__Roseburia** | -0.43378834255627 | 0.007311996018551 | 0.040943276508637 |
| **TFEQ_disinhibition_sum** | **g__Collinsella** | 0.617538379047764 | 4.67E-05 | 0.000887984951428 |
| **TFEQ_disinhibition_sum** | **g__Fusicatenibacter** | 0.519565700606822 | 0.000982453902172 | 0.009501015175819 |
| **TFEQ_disinhibition_sum** | **g__Parabacteroides** | 0.543276500476173 | 0.000511464298964 | 0.005554794801279 |
| **TFEQ_hunger_sum** | **g__Blautia** | -0.475237456810097 | 0.002954551019952 | 0.020712957676718 |
| **TFEQ_hunger_sum** | **g__Gemmiger** | -0.6262785560582 | 3.38E-05 | 0.000770182578778 |
| **TFEQ_hunger_sum** | **g__Parabacteroides** | 0.541017806719798 | 0.000545413905441 | 0.005675713453491 |
| **TFEQ_hunger_sum** | **g__Streptococcus** | -0.572305000964374 | 0.000214721845162 | 0.002750091324581 |
| **TFEQ_hunger_sum** | **TFEQ_disinhibition_sum** | 0.455169021185712 | 0.004645502001306 | 0.02946575555114 |
